# NOVA Through Two Lenses: From Nutrients to Food Processing Classification in Finland’s National Food Database

**DOI:** 10.64898/2026.10.26.26366503

**Authors:** Sherwin Nedaei Janbesaraei, Arsene Kanyamibwa, Annette Horstmann

## Abstract

Ultra-processed food (UPF) intake has been linked to adverse health outcomes. However, NOVA processing groups are hard to assign consistently, and many national food composition databases do not include them. Nutrient-based classification models could fill this gap, but their performance across countries and databases is uncertain. FoodProX is a machine-learning classifier that predicts NOVA groups and a continuous food-processing score (FPro) from nutrient composition. To test whether such models transfer across databases, we retrained it on 182,344 mostly European Open Food Facts products. We evaluated it on 45,526 held-out Open Food Facts products and, without retraining, on 2,971 NOVA-labelled foods from a US database (FNDDS). We then applied it to the Finnish national food composition database (Fineli), which has no NOVA labels. Four-class accuracy was 0.855 on held-out products and 0.763 in FNDDS, with errors concentrated at the NOVA 3/4 boundary. UPFs were identified well, with precision–recall performance of 0.940 and 0.964, respectively (on a 0–1 scale), well above the chance levels of 0.463 and 0.711. On the same items, our FPro scores ranked foods similarly to those of the originally released US-trained FoodProX (Spearman ρ = 0.81 in FNDDS and 0.79 in Fineli), although our scores were lower on average. Sodium and sugars were the strongest predictors. Low fibre and iron also pushed predictions towards NOVA 4. In Fineli, the model assigned 55.9% of the 4,092 scored items to NOVA 4. We found no earlier NOVA or FPro annotations for this database. Nutrient-based models are most reliable for detecting ultra-processing and ranking processing intensity, and less reliable for assigning all four NOVA groups. Experts also agree poorly when assigning NOVA groups, so part of this difficulty may lie in the NOVA definitions themselves.

## Introduction

Across Europe, ultra-processed foods (UPFs) now make up a large and variable share of energy intake (12.6-39.3%), with striking between-country differences (Sweden highest, Italy lowest) [1] and market shifts even in traditionally Mediterranean-diet countries (i.e., Greece) [2]. Yet classification systems remain inconsistent in distinguishing these products from less processed ones. This matters because unclear boundaries make it harder to apply processing-based dietary guidance consistently and to compare findings across studies [3–7].

High UPF intake has been associated with adverse health outcomes, including diabetes, inflammation and cognitive decline [8–11], warranting research based on a food’s processing level.[12–14][15] The NOVA classification, widely used for categorization, is based primarily on the nature and purpose of industrial processing rather than nutrient composition, although the two dimensions substantially overlap. For example, UPFs frequently share a nutritional profile with hyperpalatable foods, including combinations of sugar, fat, salt, and carbohydrates (32.9–50.2%, across 17 countries) [16]. At the same time, foods classified as UPFs can differ substantially in their nutritional profiles, including fortified products (e.g., vitamin D milk) and plant-based alternatives [17,18]. Moreover, the boundary between ultra-processed (NOVA 4) and processed (NOVA 3) foods is not always clear-cut. These discrepancies raise questions about the extent to which nutrient composition alone can reliably distinguish processing categories, and whether nutrient-based classification approaches can capture the full range of NOVA groups.

Recent reviews have highlighted NOVA’s limitations and called for improved reference methods and more consistent group assignments to strengthen research and policy [19,20]. This is particularly relevant for national food composition databases, many of which lack systematic processing classification, thus limiting research into the independent contributions of food processing and nutrient composition to health outcomes. Manual assignment of NOVA groups is labour-intensive and may introduce subjective biases, complicating comparisons across datasets. Machine learning has opened new ways to classify foods. Menichetti et al. [21] developed FoodProX, a classifier that predicts NOVA groups from nutrient composition to foods using a US database. The model also generates a continuous food processing score (FPro) for each food, ranging from 0 for a food the model is confident is unprocessed (NOVA 1) to 1 for a food it is confident is ultra-processed (NOVA 4). Unlike the four NOVA groups, FPro can rank foods by degree of predicted processing, including foods within the same group. However, it remains unclear how well such nutrient-based classifications generalize across food databases from different countries, given their differences in food composition and nutrient reporting.

To address this question, we retrained Menichetti et al.’s FoodProX classifier on predominantly European Open Food Facts data, evaluated how well it transfers to held-out European and an independent US database, and applied it to the Finnish national food composition database (Fineli), which lacks NOVA classification and a processing score.

## Methods

### Study Design

This was a methodological study based on secondary analysis of three publicly available food composition databases. We retrained the nutrient-based NOVA classifier (FoodProX) [21] on Open Food Facts [22]. Then, we evaluated it on a held-out subset of Open Food Facts and, without retraining, on an independent US database, and applied the resulting model to assign NOVA categories to the Finnish national food composition database (Fineli). For each food, we report the predicted NOVA group and the FPro score [21], calculated from the predicted probabilities of NOVA 1 and NOVA 4; the formula and its uncertainty are given in Supplementary Methods S7.

Our study design comprised three stages (Figure 1). First, we retrained the classifier on Open Food Facts. Second, transfer was assessed twice: internally, on a stratified subset of Open Food Facts held out before any model fitting (Figure 1, 2a), and externally, on the United States Department of Agriculture (USDA) Food and Nutrient Database for Dietary Studies (FNDDS) 2009–10 [23], including a head-to-head comparison against the classifier originally released by Menichetti et al. (Figure 1, 2b). Third, we applied the retrained model to Fineli. Because Fineli contains no NOVA labels, this third stage is an application rather than a validation, and no accuracy metric is computable against it; the credibility of the Fineli annotation instead rests on the two preceding validations and on agreement with the originally released classifier.

**Figure 1.**
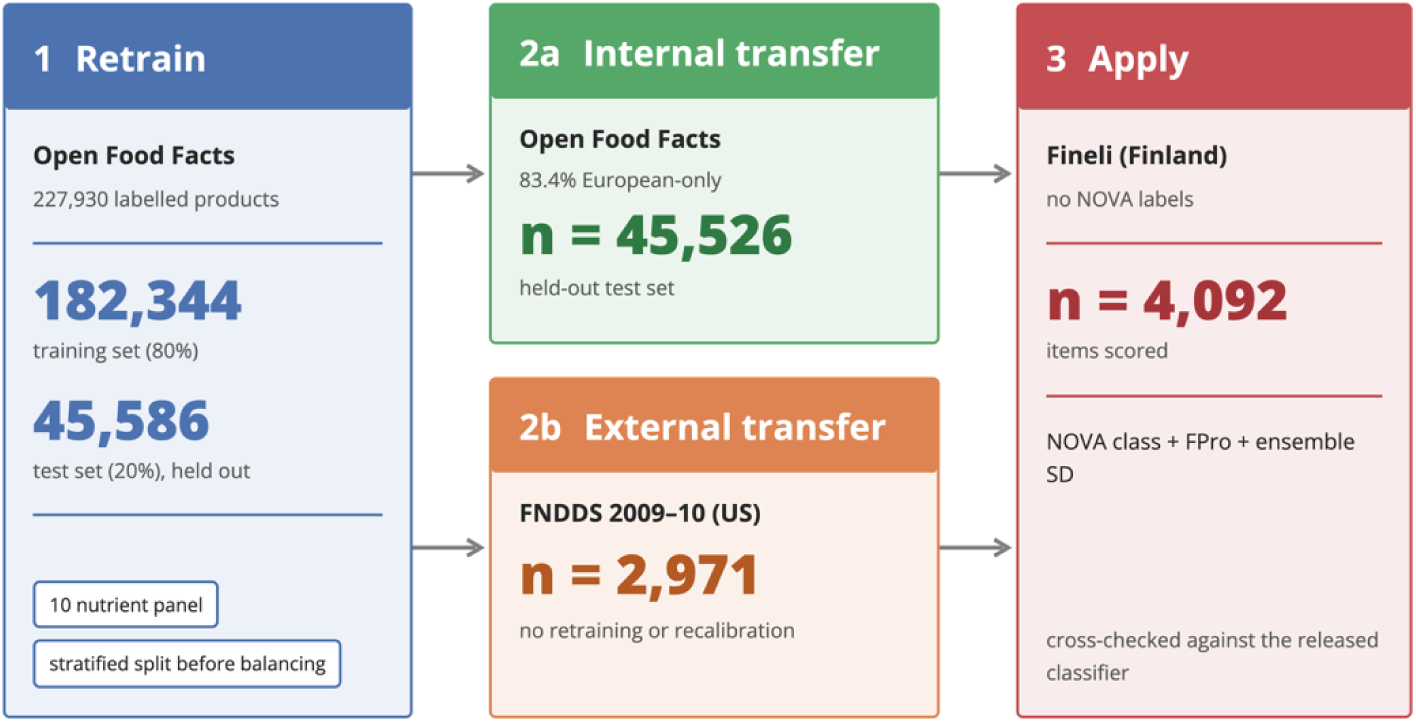
Study design. (1) The classifier was retrained on Open Food Facts (182,344 training and 45,586 test products; stratified split before class balancing). (2) Transfer was evaluated without retraining on (2a) the held-out Open Food Facts test set (n = 45,526) and (2b) the US FNDDS 2009–10 database (n = 2,971). (3) The retrained model was applied to Fineli (n = 4,092 scored). Fineli has no NOVA labels, so this stage is an application, not a validation.

### Data Sources

#### Training database

We used a subset of Open Food Facts (downloaded 15 May 2026)[22] containing 227,930 products with a NOVA group label (1–4) and complete values for all ten model nutrients. Open Food Facts is a global crowdsourced food database; however, the analytic subset was predominantly European, with 201,213 products (88.3%) listing at least one European country of sale. The subset was imbalanced across NOVA groups, with NOVA 4 being the largest and NOVA 2 the smallest class; detailed geographic and class distributions are shown in Supplementary Figure S1.

#### External validation database

We used the FNDDS 2009–10 data table distributed with the original FoodProX study [21]. It contained 7,254 foods and 106 nutrient and descriptor variables, including NOVA labels and predicted class probabilities from the released classifier [21]. Of these foods, 2,971 had a NOVA label: 2,484 assigned by NOVA researchers [24] and 487 added by the FoodProX authors [21]. The rest were unclassified (724) or mixed dishes that the original labelling split into ingredients instead of classifying (3,559). All analyses requiring reference NOVA labels used the 2,971 labelled foods. Label availability, food groups and class distributions are shown in Supplementary Figure S2.

#### Target database

We applied the classifier to Fineli, the Finnish national food composition database, comprising 4,156 food items in the file downloaded on 18 March 2026. Fineli did not include NOVA labels; it was therefore used for model application rather than accuracy evaluation. Nutrient completeness, food groups and the predicted class distributions are shown in Supplementary Figure S3.

#### Nutrient panel and cross-database harmonisation

The model uses a ten-nutrient panel expressed per 100 g (Table 1): energy (kJ), total fat, saturated fat, carbohydrate, total sugars, dietary fibre, sodium, calcium, iron and protein. All ten nutrients are reported for every one of the 227,930 labelled Open Food Facts products (100% coverage) and are available in both FNDDS and Fineli. The individual fatty-acid fractions (mono-, poly- and trans-unsaturated), which the original study’s wider panels include, are populated for fewer than 10% of labelled Open Food Facts products and were therefore excluded.

**Table 1.**
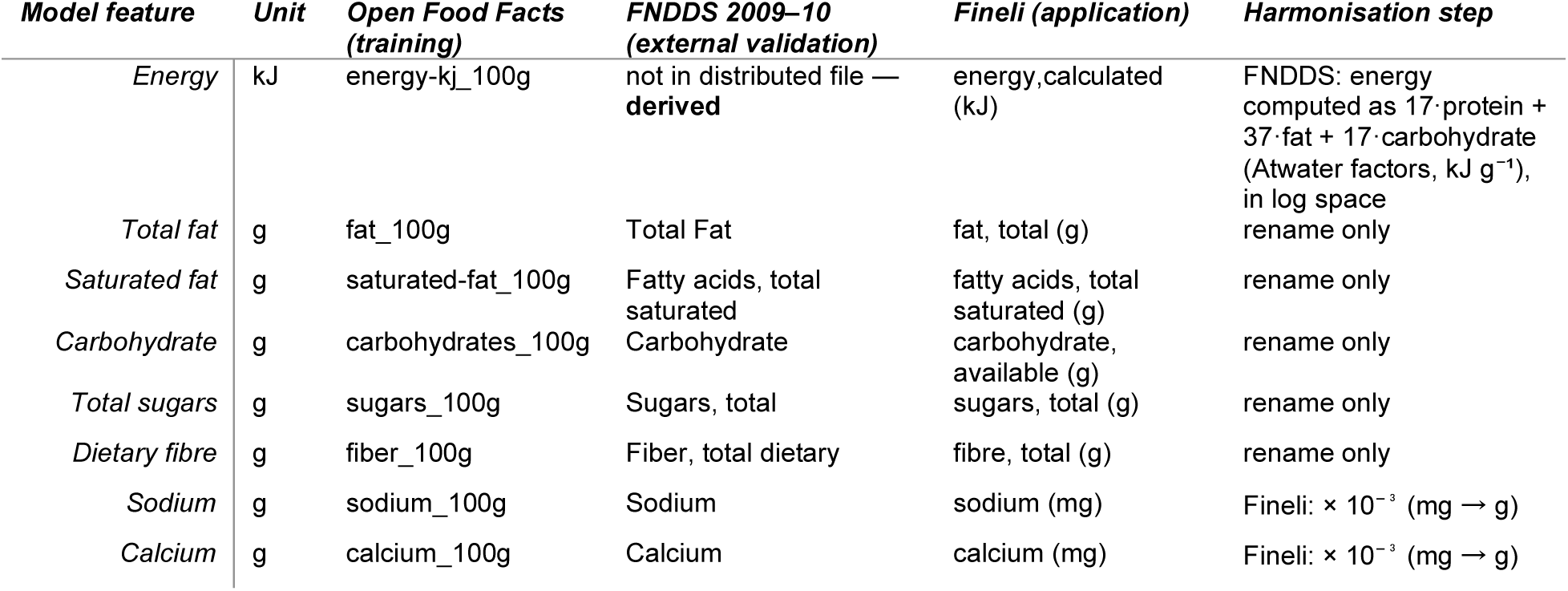
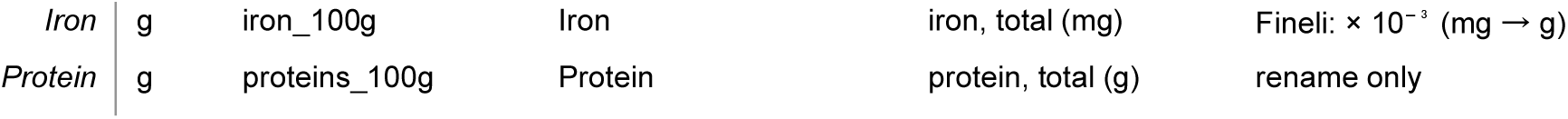
The ten-nutrient modelling panel and its source column in each database. All model features are expressed per 100 g. Open Food Facts is the training database and defines the target schema; the FNDDS and Fineli columns are mapped onto it by a dataset adapter.

Each external database was mapped onto this panel through a dataset adapter (Table 1). The distributed FNDDS table does not include energy (only nutrients measured in grams were retained [21], so energy was derived from protein, fat and carbohydrate using the Atwater-based EU labelling factors (17, 37 and 17 kJ g⁻¹) [25]. FNDDS is already natural-log transformed with all nutrients normalised to g / 100 g, so the derivation was performed in log space (exponentiate, apply Atwater factors, take the natural log). For Fineli, sodium, calcium and iron are reported in mg / 100 g and were rescaled by 10⁻³, and values recorded as below a detection limit (for example “<0.1”) were set to the threshold value rather than treated as missing. Open Food Facts and Fineli values were then cleaned (non-numeric entries set to missing, negative and incomplete rows removed), zeros replaced by 1 × 10⁻²⁰, and natural-log transformed, to match the encoding of the distributed FNDDS table. Foods retained at each step are shown in Supplementary Figure S9.

### Model Architecture and Training

We trained a nutrient-based random-forest ensemble to predict the four NOVA groups using ten nutrients per 100 g from Open Food Facts [22]. Products with valid NOVA labels and complete nutrient values were split into stratified training and held-out test sets before any modelling; negative-value removal, zero replacement and log transformation were then applied to each partition. Because some NOVA groups are much rarer than others, we added synthetic examples of the rarer groups to the training set (SMOTE) [26]. The final model averages the predictions of five sub-models, each trained on a different four-fifths of the training set. Before testing on held-out data, we estimated accuracy by training on four-fifths of the training set and testing on the remaining fifth, five times in turn (five-fold cross-validation). Following the original FoodProX code, the synthetic examples were added before this split, so the fifths used for testing also contained some of them. The held-out test set was set aside before any synthetic examples were created and contains none. The primary model was evaluated on the held-out Open Food Facts test partition and, without retraining or recalibration, on the independent FNDDS 2009–10 database. Full architecture specifications, class-balancing procedures, and sensitivity analyses are provided in Supplementary Methods S2–S4.

#### Comparison with the released FoodProX classifier

We compared our European-trained classifier with the FoodProX classifier released by Menichetti et al. (2023) [21]. That classifier was trained on US foods (FNDDS 2009–10) and uses twelve nutrients: nine of our ten (all except energy) plus vitamin C, vitamin A and cholesterol. In FNDDS, we compared our predictions with the released classifier’s predictions supplied with the data table, for the 2,971 labelled foods. In Fineli, we ran the released classifier ourselves on the 3,892 of 4,156 foods (93.6%) that report all nutrients of both panels. In both databases we report three measures of agreement: how often the two models assign the same NOVA group; how often they agree on whether a food is ultra-processed (NOVA 4 versus NOVA 1–3); and the correlation between their FPro scores (Spearman and Pearson). Details of input harmonisation, model execution and comparison metrics are provided in Supplementary Methods S6.

#### Model interpretation

We assessed the contribution of individual nutrients using permutation importance and SHAP analyses on held-out Open Food Facts data [27,28]. Detailed procedures, model configurations, sample sizes, and interpretation metrics are reported in Supplementary Methods S8. We computed permutation importance for both model configurations (primary and sensitivity); we computed SHAP values only for the sensitivity (class-balanced) configuration, whose top-ranked nutrients match the primary model (Supplementary SR4).

### Evaluation and interpretation measures

We summarised four-class classification performance using accuracy, F1 score, and recall. The reference NOVA labels are not ground truth: Open Food Facts assigns them automatically from each product’s ingredient list and additives [22] and FNDDS supplied them with the original study [21]. All three measures therefore quantify agreement with these reference labels. Accuracy is the proportion of foods assigned to their reference NOVA group; recall is the proportion of foods in a reference group correctly identified by the model; and the F1 score combines recall with precision, the proportion of predicted group assignments that are correct [29]. Permutation importance measures the reduction in classification accuracy after a nutrient is randomly shuffled, whereas SHAP values describe the contribution of each nutrient to individual model predictions [27,28]. For the binary task of distinguishing UPFs (NOVA 4) from all other foods, we report the area under the precision–recall curve (AUC-PR) and the area under the receiver-operating-characteristic curve (AUC-ROC). AUC-PR summarises the trade-off between precision and recall across thresholds and should be interpreted alongside the prevalence of UPFs, whereas AUC-ROC summarises discrimination independently of class prevalence [30,31]. We assessed agreement between the European-trained and released FoodProX models using the proportion of matching assignments and correlations between continuous FPro scores. Full definitions, formulas, and implementation details are provided in Supplementary Methods S4 and S8.

## Results

### Agreement with the released FoodProX classifier

On the 3,892 Fineli foods scorable by both nutrient panels, the European-trained classifier and the released FoodProX classifier agreed on 79.3% of four-class NOVA assignments and 82.8% of binary ultra-processed/not-ultra-processed assignments. Their continuous food-processing scores were strongly correlated (Spearman (*ρ* = 0.789); Pearson (*r* = 0.878)), although our model assigned lower scores on average (mean FPro 0.642 versus 0.704). Disagreements were concentrated at the NOVA 3/4 boundary: 504 of 807 disagreements (62.5%) involved this distinction. Most commonly where our model assigned NOVA 3, the FoodProX classifier assigned NOVA 4 (Figure 2; Table S6). In FNDDS, four-class and binary agreement were similarly 76.5% and 82.9%, respectively, with FPro scores correlating at Spearman (*ρ* = 0.813) (Supplementary Table S5). We do not report the original classifier’s FNDDS accuracy as a benchmark because FNDDS was used in its original model development and therefore does not provide an independent comparison.

**Figure 2.**
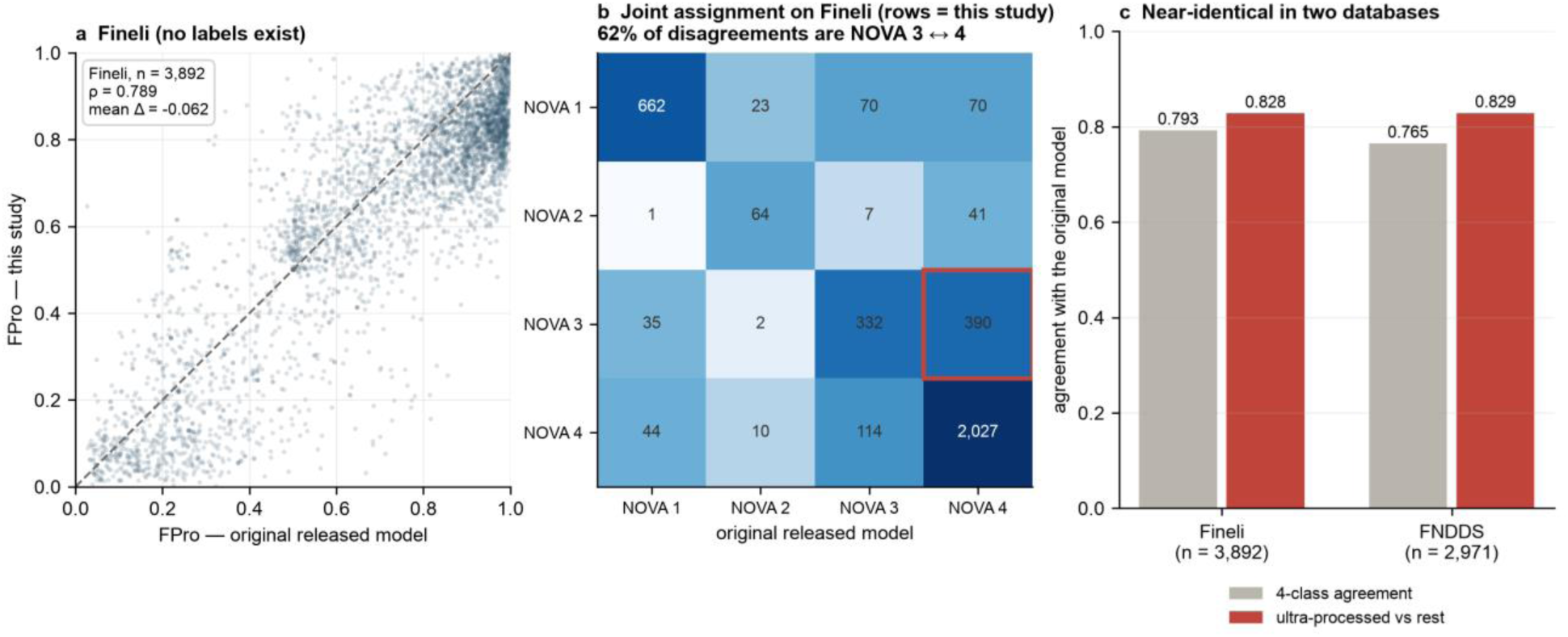
Agreement between the European-trained model and the original US-trained classifier. (a) FPro from this study against FPro from the released twelve-nutrient FoodProX classifier, for the 3,892 Fineli foods scorable by both panels; dashed line is identity. Spearman ρ = 0.789; our scores are lower on average by 0.062. (b) Joint four-class assignment of the two models on the same foods (colour on a log scale); the outlined cell (our NOVA 3, their NOVA 4) is the single largest disagreement (390 foods, 48.3% of disagreements; with the reverse cell, 114, the 3/4 boundary accounts for 504 of 807, 62.5%). (c) Class agreement between the two models on the four-class task and on the ultra-processed / not distinction, in Fineli (n = 3,892) and FNDDS (n = 2,971); UPF agreement is near-identical in the two databases (82.8% and 82.9%).

### Nutrient composition separates the poles of the NOVA scale, not its middle

On the held-out Open Food Facts test set (n=45,526), four-class accuracy was 0.855. The classifier performed well for NOVA 1, NOVA 2, and NOVA 4 (F1 = 0.92, 0.91, and 0.87, respectively), but performance was weaker for NOVA 3 (F1 = 0.77; Figure 3a; Supplementary Table S2). Misclassifications were concentrated at the processed/ultra-processed boundary: NOVA 3↔4 errors accounted for 71.6% of all errors in the held-out test set (Figure 3b). Thus, nutrient composition differentiated the less processed and ultra-processed ends of the NOVA scale more reliably than the intermediate processed category.

**Figure 3.**
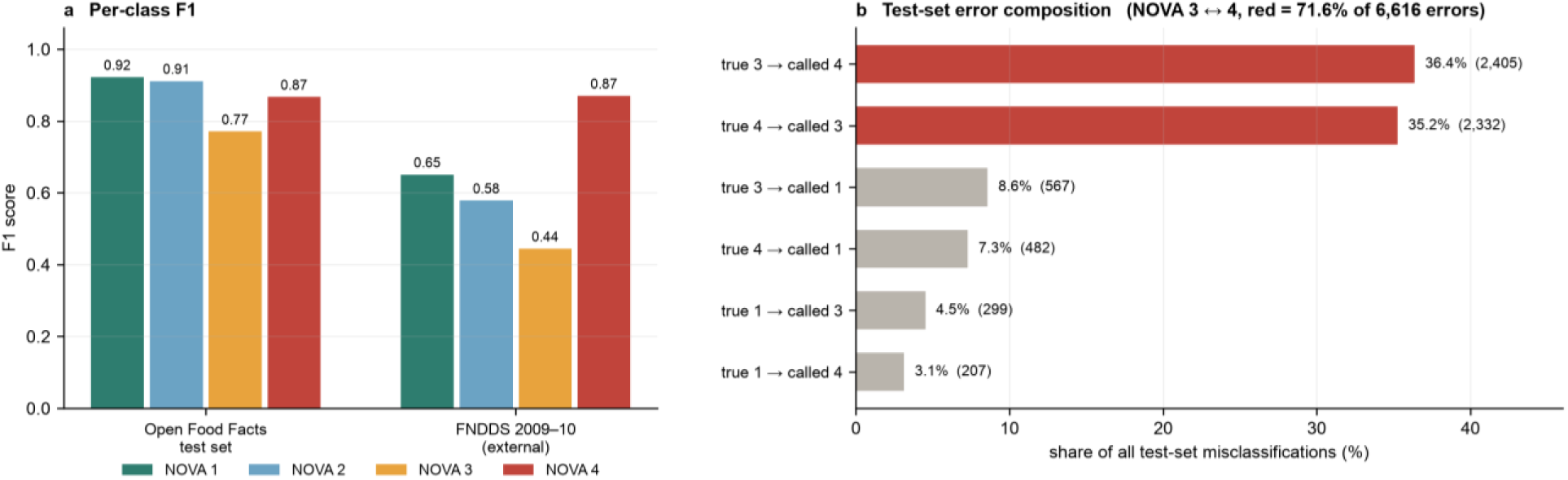
Nutrient composition separates the poles of the NOVA scale, not its middle. (a) Per-class F1 score for the primary configuration on the Open Food Facts test set (n = 45,526) and on FNDDS (n = 2,971). In Europe, NOVA 1, 2, and 4 are recovered well (F1 ≥ 0.87), and NOVA 3 is the weakest class (F1 = 0.77). Under transfer to the United States, F1 falls for NOVA 1–3 (to 0.65, 0.58 and 0.44) and is maintained only for NOVA 4 (0.87); NOVA 3 shows the largest loss of recall (0.76 → 0.44), and the NOVA 2 estimate rests on 54 foods. (b) Composition of the 6,616 held-out misclassifications: the six most common error types as a share of all errors. Bars for the NOVA 3 ↔ 4 confusion are red; together they account for 71.6% of all errors, whereas the next most common errors, NOVA 3 → 1 and NOVA 4 → 1, account for 8.6% and 7.3%.

Nutrient composition therefore carries a strong signal separating MPFs from UPFs and a weak one for the intermediate “processed” category. This same boundary is also where recall degrades most when the model is transferred to the US food supply (NOVA 3 recall 0.76 → 0.44; Figure 3a, Table S4). It is likewise where our model and the independently trained original classifier disagree most (Figure 2b), and where the five sub-models disagree most about the unlabelled Finnish foods (Supplementary Figure S7).

### Discrimination of UPFs

Because the practically consequential question is usually whether a food is ultra-processed, we also evaluated the binary task of separating NOVA 4 from NOVA groups 1–3 across all decision thresholds (Figure 4, Table S3). On the Open Food Facts test set, the AUC-PR curve was 0.9399, compared with a prevalence baseline of 0.463 (AUC-ROC = 0.9513). In FNDDS the AUC-PR was 0.9635 against a baseline of 0.7109 (AUC-ROC=0.9191). The higher FNDDS figure reflects a higher prevalence of UPFs in that database, not better discrimination: measured against its own baseline, the European held-out result is the stronger of the two. The contrast with the four-class results is informative: the model is only moderately accurate at reconstructing the full NOVA taxonomy but is a strong detector of ultra-processing specifically.

**Figure 4.**
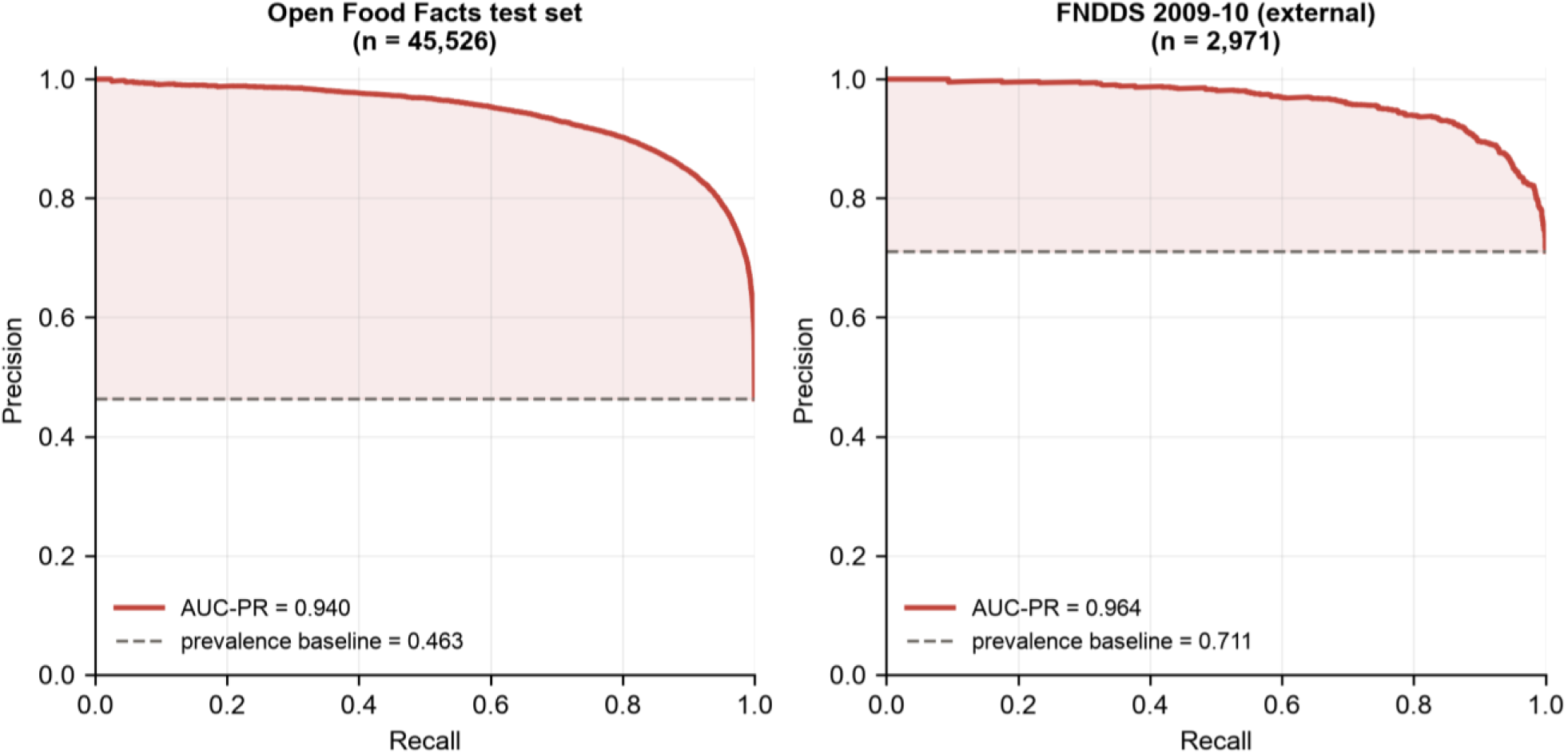
Detection of UPFs (NOVA 4 vs NOVA 1–3). Precision–recall curves for the primary classifier in the held-out Open Food Facts test set and FNDDS 2009–10. Dashed horizontal lines indicate the prevalence of UPFs in each dataset.

When applied without retraining to FNDDS, four-class accuracy declined to 0.763, driven mainly by poorer recovery of NOVA 3 (recall 0.76 → 0.44), whereas recall for NOVA 1 and NOVA 4 changed little (0.95 → 0.88 and 0.86 → 0.82; Figure 3a). F1 fell for NOVA 1–3 (0.92 → 0.65, 0.91 → 0.58 and 0.77 → 0.44) but not for NOVA 4 (0.87 in both databases). For NOVA 1, the loss was almost entirely in precision (0.90 → 0.52), because 271 US NOVA 3 and NOVA 4 foods (9.1% of FNDDS) were assigned to NOVA 1; NOVA 2 lost both precision and recall and rests on only 54 foods. The unchanged NOVA 4 F1 reflects lower recall offset by higher precision (0.87 → 0.93), consistent with its higher prevalence in FNDDS (71% vs 46%). Discrimination of UPFs remained high (Figure 4; Supplementary Tables S2–S4).

### Nutrient drivers of the classification

Permutation importance (Table S7, Figure 5) identified sodium as by far the dominant predictor in both configurations. In the primary configuration (20,000 held-out products, 5 repeats), permuting it cost 15.5 accuracy points, or 18.1% of baseline accuracy. Total sugars ranked second (8.6 points; 10.1%) and protein third (6.1 points; 7.2%); every other nutrient contributed under 5 points. The variables most associated with energy density contributed least: total energy ranked ninth of ten (3.3 points; 3.9%) and saturated fat last (2.0 points; 2.4%). Permutation importance and SHAP agree once they answer the same question: on the class-balanced model used for SHAP, both rank sodium first across all four classes, and permutation importance ranks sugars second. Sugars led only for the probability of NOVA 4, with sodium a close second (Supplementary Figure S5a). Sodium dominates overall accuracy because it separates NOVA 1 from NOVA 3, which together make up about half of the foods: low sodium pushes a food towards NOVA 1 and high sodium towards NOVA 3 (Supplementary SR4).

**Figure 5.**
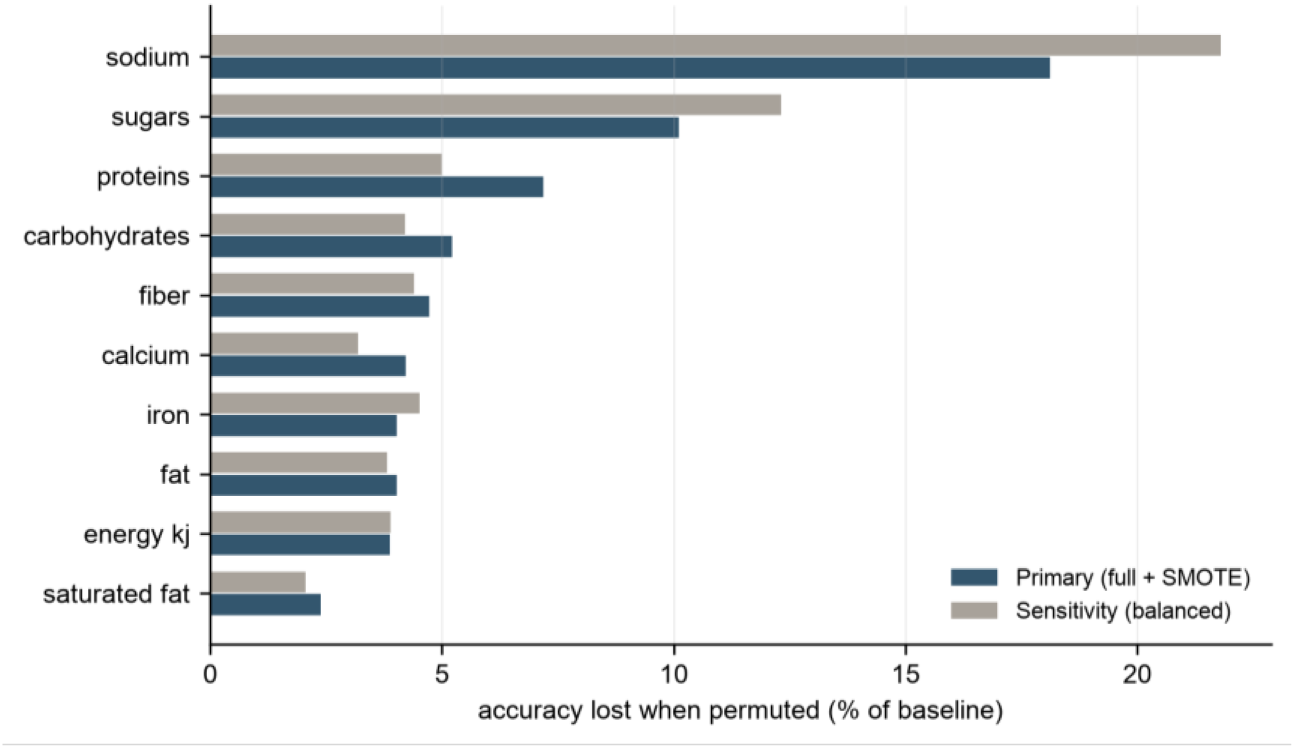
Permutation importance. Loss in test-set accuracy (as a percentage of baseline accuracy) when each nutrient is randomly permuted, for both model configurations; computed on 20,000 (primary) and 2,000 (sensitivity) test-set products, with 5 and 2 repeats. Sodium and total sugars occupy the top two positions in both configurations; total energy and saturated fat contribute least.

### Application to the Finnish national food composition database

#### Class and score distribution

Of 4,156 Fineli items, 4,092 (98.5%) had complete values for the ten-nutrient panel and were assigned a NOVA class and FPro score. The model assigned 20.9% of scored items to NOVA 1, 2.8% to NOVA 2, 20.4% to NOVA 3, and 55.9% to NOVA 4. Mean FPro was 0.642 (median 0.738; SD 0.269), and its distribution was bimodal (Figure 6; Supplementary Figure S6), consistent with the model’s tendency to separate the poles of the scale confidently.

**Figure 6.**
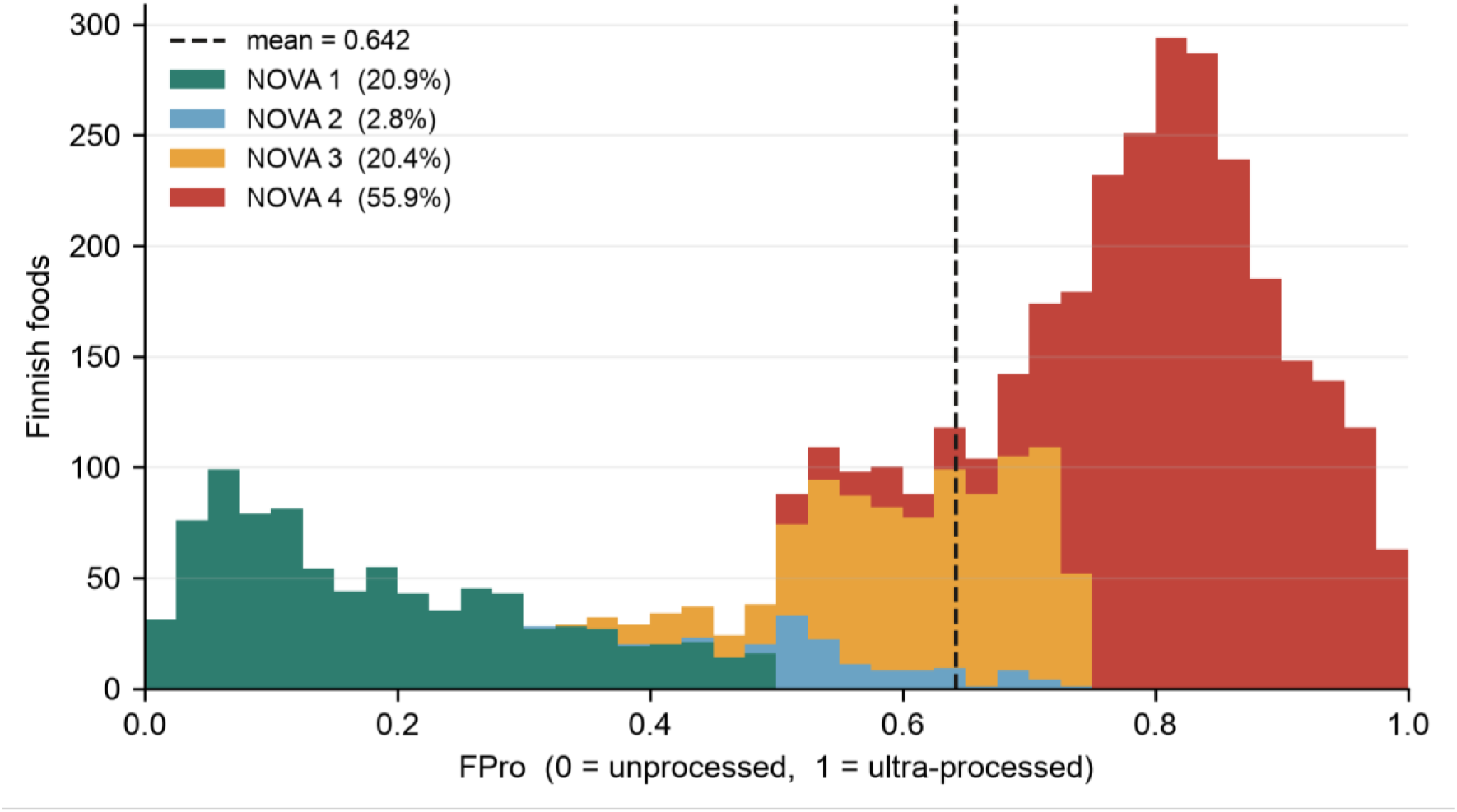
FPro distribution of the Finnish food composition database. Stacked histogram of FPro for the 4,092 scored Fineli items, coloured by the NOVA class the model assigned; dashed line is the mean (0.642). The distribution is strongly bimodal, consistent with a model that separates the poles of the scale confidently and is less decisive for intermediate foods. These are model predictions: Fineli carries no NOVA labels, and the proportions describe a food composition database, not Finnish dietary intake.

#### Face validity

Because Fineli has no reference NOVA labels, its predicted classifications cannot be evaluated against a ground truth. Their credibility therefore rests on the held-out and external validation results, agreement with the independently trained released FoodProX classifier, and qualitative face validity within the Finnish database. Food groups derived from item names showed the expected ordering of median FPro scores: fruits and berries were lowest (0.211), followed by vegetables and legumes (0.246), whereas composite dishes and ready meals (0.797) and sweets and snacks (0.867) were highest (Supplementary Figure S8). In addition, 64.8% of fats and oils were assigned to NOVA 2, compared with 2.8% of all Fineli items, consistent with NOVA’s category of processed culinary ingredients.

FPro also generally tracked formulation and preparation within food families (Figure 7). Scores started from 0.02 for raw chicken breast to 0.95–0.97 for nuggets, industrial meatballs, and cold cuts; from 0.06 for salmon fillet to 0.90 for fish fingers; and from 0.005 for rolled oats to 0.92–0.99 for oat biscuits. However, the ladders also show the limits of a nutrient-only score. Across all Fineli items named “Yoghurt, flavoured” (n = 51) and “Ice cream” (n = 27), the two families overlapped almost completely (median FPro 0.87 and 0.96; both ranging from 0.52 to 0.99), and home-made and industrial oat biscuits had the same median (0.92; n = 3 and 4). Single-item contrasts therefore depend on the recipe chosen: the ice cream in Figure 7 lies near the bottom of its family (4th percentile) and the flavoured yoghurt near the top (98th percentile). These cases show that FPro captures nutrient patterns associated with processing, rather than directly observing ingredient lists, industrial formulation, or processing operations.

**Figure 7.**
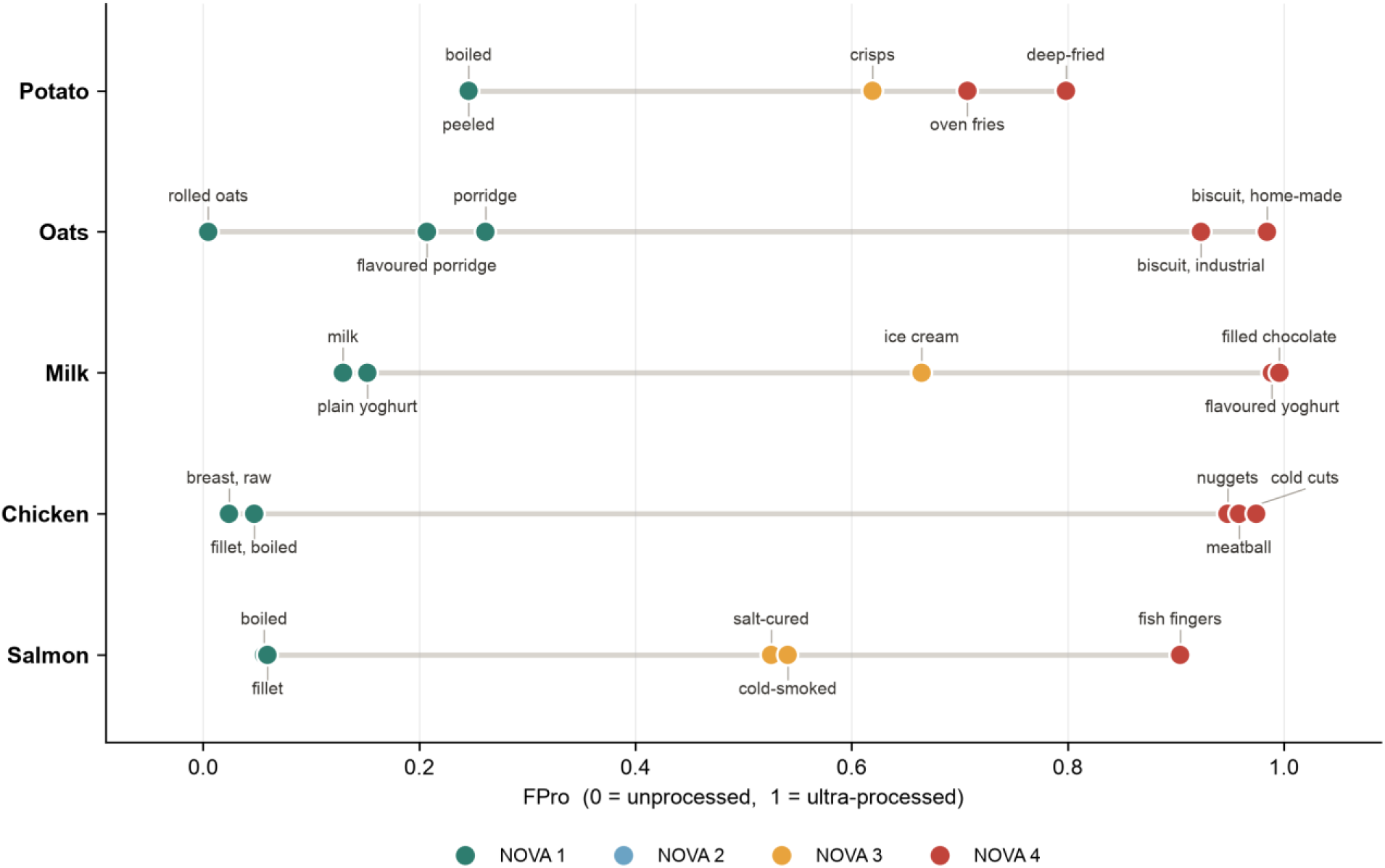
Processing ladders within Finnish food families. For five base ingredients, named Fineli items are placed on a common FPro axis and coloured by assigned NOVA class; each label is joined to its item by a short line. Items illustrate preparation steps and are not representative of their food families. Within each family the score tracks preparation and industrial formulation, for example potato from 0.25 (peeled) to 0.80 (deep-fried) and chicken from 0.02 (raw breast) to 0.95–0.97 (nuggets, industrial meatballs, cold cuts). This reproduces, in a Finnish database and with an independently trained model, the within-family gradation the original study showed for onion preparations.

## Discussion

We retrained the FoodProX approach of Menichetti et al. (2023) [21] on European Open Food Facts data, evaluated it on held-out Open Food Facts products and, without retraining, on the US FNDDS database, and then applied it to the Finnish Food Composition Database (Fineli). To our knowledge, this work also provides the first NOVA and continuous food-processing-score (FPro) annotations for Fineli. Nutrient-based models reliably identified UPFs across databases, but their ability to reconstruct the full NOVA groups was limited, particularly at the processed/ultra-processed boundary.

### Model classifier performance

On Open Food Facts products, the model had never seen, accuracy was 0.855 (Table S2), about six percentage points below the cross-validation estimate (0.911; Table S1). That estimate is optimistic for two reasons. First, following the original FoodProX code, synthetic examples of the rarer NOVA groups were created before the training data were divided into fifths, so some synthetic training foods were blended from foods later used to test the model. Second, the estimate gave all four NOVA groups equal weight, whereas the held-out products keep their natural proportions; weighting the groups equally, held-out accuracy was 0.880, which closes about half of the gap. By contrast, the model identified UPFs far better than chance (AUC-PR 0.940 against a baseline of 0.463).

These results indicate that the classifier substantially outperforms the baseline and achieves higher precision-recall performance in the target NOVA classes. (Table S3, Figure 4). This performance also held when we compared the European test set (Open Food Facts (AUC-PR = 0.9399) with FNDDS (AUC-PR = 0.9635) (Table S3, Figure 4). Interestingly, the classifier’s errors were not distributed evenly across the NOVA classification but concentrated at a single boundary. Confusion between processed foods (NOVA 3) and UPFs (NOVA 4) accounted for 71.6% of all misclassifications in the test set, whereas confusion between the two ends of the scale was much less common (NOVA 1 → 4, 3.1%; NOVA 4 → 1, 7.3% of errors). The same pattern emerged across the other analyses. When transferred to the US food supply, recall for NOVA 3 fell from 0.76 to 0.44, whereas recall for NOVA 1 and NOVA 4 changed little (0.95 to 0.88 and 0.86 to 0.82; Figure 3a, Table S4). Among Finnish foods for which our European-trained model and the independently trained original classifier disagreed, 62.5% of disagreements involved the NOVA 3/4 distinction (Figure 2b). Similarly, all models’ disagreement on NOVA prediction (ensemble uncertainty) in Fineli data was greatest for foods with intermediate processing scores (Figure S7). The convergence of these findings across two continents, three databases, and two independently trained models suggests that the NOVA 3/4 boundary poses a reproducible challenge for nutrient-only classification rather than reflecting an idiosyncrasy of a single training set. This difficulty follows from how NOVA defines the two groups. Processed foods (NOVA 3) are made by adding salt, sugar or oil to MPFs, as in cheese, cured meats or freshly baked bread, whereas UPFs (NOVA 4) are defined by industrial ingredients and cosmetic additives, such as emulsifiers, flavourings and colourings, with no domestic culinary equivalent [32]. These additives are used in small amounts and barely change a food’s nutrient profile, and both groups tend to be high in salt, sugar or fat: a bakery loaf and a packaged loaf with emulsifiers can have almost the same nutrients yet fall on opposite sides of the boundary. Our model reflects this, as high sodium pushes foods towards both NOVA 3 and NOVA 4 (Supplementary SR4). These results corroborate evidence from experts and the public about the occasional difficulty of distinguishing NOVA groups 3 and 4 [6,7,33,34].

Hence, our work shows that FoodProX can be generalized across food-composition systems and that its performance is task-dependent. Nutrient composition appears to provide a robust signal for distinguishing UPFs from less processed foods, whereas it provides a weaker basis for resolving the intermediate NOVA 3 category (and, under transfer, NOVA 2). Accordingly, the model should be interpreted as a strong detector of ultra-processing and a useful ranking tool for processing intensity, rather than as an equally reliable substitute for expert assignment across all four NOVA categories.

These findings have implications beyond food-composition database annotation. For nutritional epidemiology, a reproducible nutrient-based UPF indicator could improve the coverage and comparability of exposure assessment when ingredient lists, or expert NOVA labels are incomplete. Moreover, as argued by Romero Ferreiro et al. [35] nutrient information and the NOVA labels can be combined to better inform nutrient quality. Our results also illustrate how structured nutrient data can complement ingredient-based and language-model approaches [36,37], particularly when product descriptions are unavailable or inconsistently recorded. However, these applications should treat model-derived annotations as probabilistic measurements rather than ground-truth labels. Linking food processing to disease prevalence is not a simple or straightforward task, and caution is warranted [38,39]. Hence, we should preserve uncertainty around foods whose nutrient profiles do not clearly distinguish between adjacent NOVA categories.

### What FPro measures across food databases

Having established that nutrient composition supports strong UPF detection but only partial resolution of the NOVA scale, we next ask how the resulting FPro scores transfer across food databases. When applied to FNDDS without recalibration, NOVA assignment accuracy declined by approximately nine percentage points, from 0.855 to 0.763 (Table S2), whereas discrimination of UPFs remained comparatively strong (AUC-ROC = 0.919) (Table S3). Transfer also altered the error distribution: in the European evaluation, misclassifications occurred predominantly between adjacent NOVA classes, whereas in the US data, a substantial proportion of true NOVA 3 and NOVA 4 foods were assigned to NOVA 1 (Figure S4).

More importantly, transfer affected absolute FPro values more than ranking (relative order). The model systematically assigned lower FPro values to US foods than did the original classifier [21], with mean scores of 0.678 and 0.774, respectively, for the same items (Table S5). A similar downward offset was observed in Fineli, with corresponding means of 0.642 and 0.704 (Figure 2, Table S6). These differences could reflect genuine variation between food supplies, differences in reference labels, or the reduced ten-nutrient input panel. The present analyses cannot distinguish among these explanations. Nonetheless, the relative ordering of foods was considerably more stable, with Spearman correlations of 0.79–0.81 between the two models (Table S5, S6). This establishes a key distinction: absolute FPro values should not be assumed directly comparable across models or databases, whereas the rank ordering of foods by processing intensity is much more robust. These results might also reflect differences in regulatory approaches between the US Food and Drug Administration (FDA), the European Food Safety Authority (EFSA), the World Health Organization (WHO), and the Food and Agriculture Organization (FAO). Despite coordination efforts and similarities in risk assessment procedures, differences remain in which additives are recognized as safe [40,41] which could affect the nutrient composition of the same products. This highlights the importance of applying FoodProX across different nutrient databases.

In Fineli, the classifier assigned 55.9% of records to NOVA 4 and produced a strongly bimodal FPro distribution; ensemble uncertainty was low for most records, with 80.4% having uncertainty below the threshold (*σ*_FPro_ < 0.02) (Supplementary Figure S7). An important limitation is the absence of independently assigned NOVA reference labels for Fineli. Consequently, the accuracy of the resulting annotations cannot be directly established, and agreement between classifiers or low ensemble uncertainty should not be interpreted as independent validation. For example, the model assigned 64.8% of fats and oils to NOVA 2, corresponding to NOVA’s category of processed culinary ingredients, and generated plausible within-family processing gradients: FPro increased from 0.02 for raw chicken breast to 0.95–0.97 for nuggets and cold cuts, and from 0.005 for rolled oats to 0.92–0.99 for oat biscuits (Figure 7). These gradients should not, however, be interpreted as direct measurements of industrial processing. Although processing can alter nutrient profiles, industrial processing is a highly complex set of operations that includes ingredient choices, additive functions, and technological steps [42] that nutrients alone may not fully capture. Our model therefore ranks foods by nutrient patterns associated with processing, not by observed processing operations or ingredient list features [21]. This approach rests on an assumption supported by earlier work: in unprocessed foods, each nutrient varies within a characteristic natural range, and processing tends to move nutrient concentrations outside that range, for example through sugar added to a flavoured yoghurt or fibre lost when grain is refined [21,43]. However, the overlap between flavoured yoghurts and ice creams illustrates this limitation: both families span the same FPro range (0.52–0.99), so single-item contrasts such as the 0.99 and 0.67 in Figure 7 reflect individual recipes rather than a systematic difference between food types, because FPro reflects nutrient content rather than formulation or processing steps directly. Finally, the Fineli class proportions describe the database, not the Finnish diet. Each item counts once, however often it is eaten, so a rarely eaten ready meal weighs as much as rye bread or milk, and the database’s coverage of composite dishes and branded products shapes the proportions further. The 55.9% of Fineli items assigned to NOVA 4 is therefore a share of foods listed, not of foods eaten, and cannot be compared with dietary estimates such as the UPF share of the diet in Finland (29.6%) [1] or the United States (53.4%) [44]. It is closer in kind to studies of the food supply, which document the dominance of UPFs in food systems and retail [45–49]. Estimating UPF intake in Finland would require linking the annotation to dietary intake data with consumption weights.

### Processing or Nutrients?

The NOVA classification was driven by a narrow set of nutrients. Permuting sodium reduced accuracy by 18.1% relative to baseline, and permuting total sugars by 10.1%, whereas total energy and saturated fat contributed least (3.9% and 2.4%, respectively; Figure 5). Sugar was the dominant signal for the UPF probability, sodium for the unprocessed and processed probabilities, and protein for culinary ingredients (Fig S5a). We found that two of the three components (sugar and salt) do indeed carry most of the predictive signal, but fat does not (Figure 5, S5). Most importantly, across the foods examined, the rank correlation with the UPF probability was +0.87 for sugars and +0.84 for sodium (Table S8). These findings demonstrate that the classifier relies strongly on nutrient characteristics associated with NOVA categories, rather than directly identifying the industrial processes or ingredients used in food production. The dominance of sugar and sodium is consistent with the substantial, though incomplete, overlap between ultra-processed and hyper-palatable foods, indicating that nutrient profiles capture an important component of ultra-processing without being synonymous with it [16,50]. In a nationally representative US study, UPFs supplied 89.7% of energy from added sugars, and greater processing has been linked to higher glycaemic responses [24,51]. Similarly, higher UPF intake has been associated with incident hypertension [52], consistent with sodium reduction being an established strategy for hypertension prevention [53–57]. These findings provide a broader nutritional context for the patterns identified by our classifier, but do not establish that its predictions capture processing-related health risks independently of nutrient composition.

Lastly, iron and dietary fibre showed negative associations with predicted UPF probability, with rank correlations of −0.59 and −0.57, respectively (Table S8). These associations indicate that the model does not rely exclusively on high sugar, sodium or fat content, but also captures other differences in nutrient composition across NOVA groups. Instead, it seems to capture features consistent with degradation of the food matrix, such as lower fiber and mineral density, which are reflecting compositional changes associated with food processing [58,59].

In our view, the NOVA Classification is reasonably contested, with good arguments for and against [60,61]. On the one hand, not all UPFs are created equal, with a lot of variability between items and a lack of high-quality evidence for plant-based alternatives [62]. Nevertheless, UPFs and processed diets are also less likely to provide a sufficient amount of micronutrients [63], and when they do, they might cause higher energy intake [64]. One UPF study also found that vegan youth (16-24 years old) in Sweden consumed more energy and nutrients like salt [65], whereas a UK-based RCT observed that participants lost more weight on an MPF diet versus a UPF diet that matches national recommendations [12]. NOVA is also hard to apply consistently by hand: French nutrition specialists agreed poorly when classifying the same foods, even when ingredient information was given (Fleiss’ κ 0.32 and 0.34, respectively, where 1 indicates full agreement) [7]. Their results corroborate recent evidence that suggests that NOVA may not the best tool to evaluate food processing [66], but without more ingredient information from the food industry, NOVA remains the easiest to apply. This limits what any evaluation of our annotation can show. However, one could test the Fineli annotation directly by having nutrition professionals, each assign a random sample of Fineli foods to NOVA groups without seeing the model’s predictions. The model’s agreement with such labels should then be judged against how well the professionals agree with one another.

Thus, whether we use NOVA or other methods to identify processing levels, they provide information on national dietary consumption patterns, the utility of functional health foods [70], and a link to global food system transformation [19] driven by corporate influence [67–69].

## Conclusion

We retrained a nutrient-based NOVA classifier (FoodProX), tested it on European and US food databases and used it to annotate the Finnish national food composition database (Fineli). It consistently ranked foods by predicted processing intensity and detected UPFs from nutrient information but could not equally resolve all four NOVA groups. Once linked to dietary intake data, the Fineli annotation could support future nutritional epidemiological research in Finland. Our findings delineate the theoretical and practical boundaries of nutrient-only NOVA proxies. Rather than serving as universal class classifiers, these models function most reliably as binary detectors of ultra-processing and as continuous ranking tools for processing intensity. The reduced accuracy at the NOVA 3/4 boundary underscores that nutrient profiles alone cannot fully encode the procedural and additive criteria central to NOVA’s definition of ultra-processing. To understand how food relates to disease risk across the spectrum from MPFs to UPFs, we need to understand its chemical complexity [70]. A key improvement for FoodProX would be to train it on food composition databases that include detailed additive and ingredient information (e.g., E170: calcium carbonate). The model’s agreement with such labels should therefore be judged against how well the professionals agree with one another. Consequently, model-derived annotations should be treated as probabilistic measurements, with key boundaries interpreted as provisional and used alongside, not in place of, expert judgment and ingredient-based methods.

## Data Availability

Code availability. Analysis code, trained-model outputs, and instructions to reproduce the reported analyses are available at https://gitlab.com/obrainlab-group/nutrients-to-processing-in-fineli.

## Acknowledgments

SNJ was supported by the Finnish National Agency for Education (EDUFI). SNJ and AH were also supported by the Kristian Gerhard Jebsen Foundation within the framework of the Centre for Obesity Research (CORA). AK was supported by funding from the Jenny and Antti Wihuri Foundation and the Instrumentarium Science Foundation. We sincerely thank Giulia Meninchetti for her feedback on the analysis plan.

## Author contributions

**SNJ:** Conceptualization, Methodology, Writing original draft, Formal analysis, Investigation, Data curation, Software, Visualization, Resources, Writing-Review & editing, Funding acquisition, Project administration. **AK:** Conceptualization, Methodology, Writing original draft, Investigation, Data curation, Resources, Writing-Review & editing, Funding acquisition, Project administration. **AH:** Conceptualization, Methodology, Supervision, Project administration, Funding acquisition, Writing-Review & editing.

## Ethical Statements

This study was conducted in compliance with the principles of the Declaration of Helsinki. This study is a secondary analysis of publicly available food composition databases (Open Food Facts, USDA FNDDS 2009–10 and the Finnish Fineli database). It involved no human participants and no personal, identifiable or sensitive data. Ethical approval and informed consent were therefore not required.

## Ethics declarations

The authors declare no competing commercial or financial interests.

## Declaration of Generative AI and AI-assisted technologies in the writing process

During the writing of this work, SNJ and AK used Claude AI Sonnet 5.5 to improve readability and language. AI was also used to draw Figure 1 of manuscript. Afterward, SNJ and AK reviewed and edited the content as needed and take full responsibility for the publication’s content. The same AI model was also used to add comments to the code available in the repository.

