## Supplementary Material for "NOVA Through Two Lenses: From Nutrients to Food Processing Classification in Finland’s National Food Database"

#### Supplementary Methods

##### S1. Data sources, preprocessing, and harmonisation

We used an extract of Open Food Facts containing 227,930 products with a NOVA group label (1–4) and complete values for all ten nutrients of the modelling panel drawn from the Open Food Facts database export downloaded on 15 May 2026. Open Food Facts is a global crowdsourced database, but this labelled extract is predominantly European. Of this extract, 201,213 products (88.3%) list at least one European country of sale, 190,156 (83.4%) list only European countries, and 26,717 (11.7%) list no European country, most of them the United States or Canada. Counting each product once for every country it lists, the extract is strongly skewed toward France (116,782 products, 51.2%; 88,880 sold only in France), followed by Germany (39,772; 17.4%), the United States (22,257; 9.8%), the United Kingdom (16,350; 7.2%), Spain (15,523; 6.8%) and Italy (11,496; 5.0%). The NOVA class distribution of the extract was 49,912 (NOVA 1), 8,715 (NOVA 2), 63,602 (NOVA 3), and 105,701 (NOVA 4).

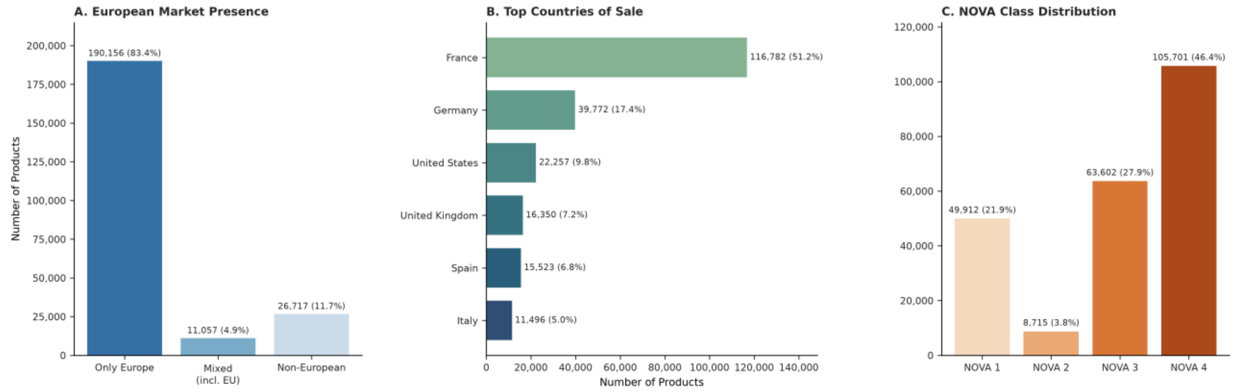


**Figure S1**. **Overview of the labelled Open Food Facts dataset extract (N = 227,930).** (A) Product distribution by geographic market presence, illustrating the dataset's predominantly European focus. (B) The six most frequently listed countries of sale; a product can list several. France is listed for 51.2% of products and is the only country of sale for 39.0%. (C) Distribution of products across the four NOVA processing categories, ranging from unprocessed/minimally processed (NOVA 1) to ultra-processed (NOVA 4).

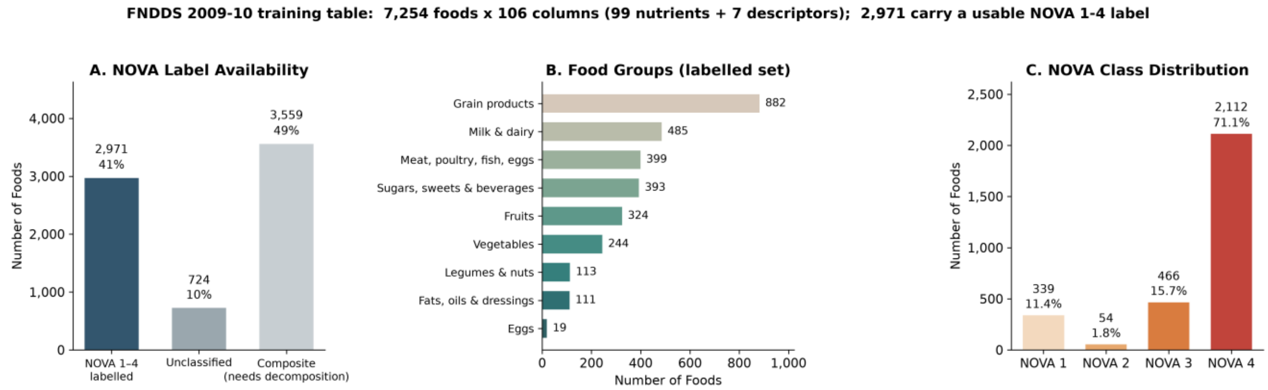


**Figure S2. Summary of the FNDDS 2009–10 external validation dataset (N = 7,254 foods).** (A) Availability of NOVA labels: foods with a NOVA 1–4 label, foods without one (unclassified), and composite dishes, which carry no single NOVA label in the source table. (B) The 2,971 NOVA-labelled foods by USDA food group, taken from the first digit of the food code. (C) Distribution of the labelled foods across the four NOVA groups.

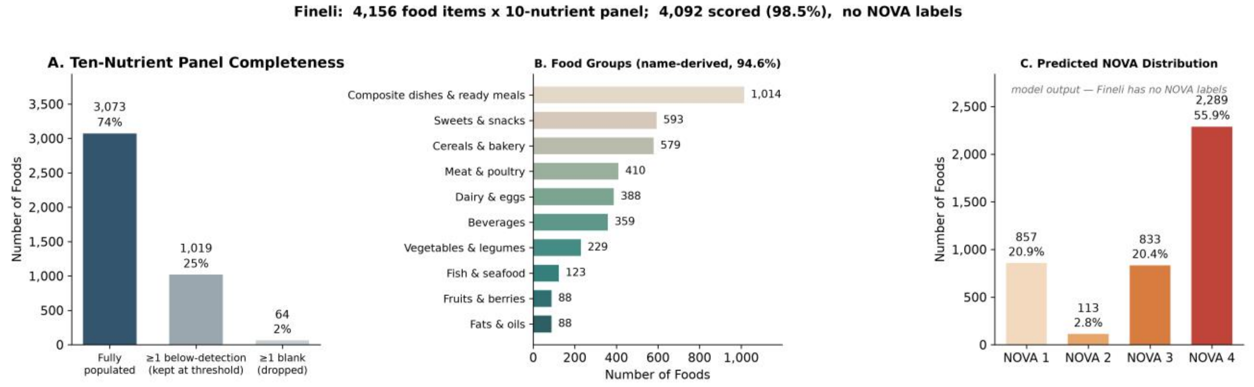

**Figure S3. Summary of the Fineli dataset (N = 4,156 food items**). (A) Completeness of the 10-nutrient panel. The majority of items (74%) are fully populated, 25% contain at least one below-detection value (kept at the threshold), and 2% were dropped due to blank entries, resulting in 4,092 (98.5%) successfully scored foods. Breakdown of items by name-derived food groups, with composite dishes and ready meals representing the largest category (1,014 items). (C) The model-predicted NOVA distribution, as the original Fineli database lacks ground-truth NOVA labels. Over half of the scored foods (55.9%) are predicted to be ultra-processed (NOVA 4).

**Preprocessing and dataset split.** Products were retained when they had a valid NOVA label and non-missing values for all ten nutrient features. Duplicate product codes were removed by retaining the first entry; nutrient fields were converted to numeric values, and nutrient fields were converted to numeric values. Before class balancing or model fitting, the labelled Open Food Facts products with complete nutrient panels were split 80/20 into stratified training and held-out test partitions using a fixed random seed (182,344 training and 45,586 test products). Rows with negative nutrient values were then removed from each partition (208 training, 60 test), zeros were replaced with $1 \times 10^{-20}$, and nutrient values were natural-log transformed. The final training matrix contained 182,136 products and the held-out test matrix 45,526 products.

**Cross-database harmonisation.** The ten-nutrient panel and source-column mappings are presented in Table 1 of the main text. FNDDS energy was derived from protein, fat, and carbohydrate using Atwater factors, while sodium, calcium, and iron in Fineli were converted from mg/100 g to g/100 g. Values reported below a Fineli detection threshold were retained at their threshold rather than treated as missing.

#### S2. Model architecture and ensemble training

In alignment with the actual models distributed by the primary authors, we implemented a scikit-learn random forest ensemble consisting of 1,800 trees with max features='sqrt' and max depth=260. Although the original publication described a setup using 200 trees and a depth cap of 420, inspection of the three publicly shared models confirmed they were configured with 1,800 trees and a depth threshold of 260. We chose to match these public artifacts to maintain fidelity to the empirical findings. Crucially, this depth constraint was non-binding in practice, as trees attained peak depths of only 40 in our retraining and 29 in the reference model, leaving all decision pathways untruncated.

To handle class skew, the Synthetic Minority Over-sampling Technique (SMOTE) was executed on the training split prior to ensemble generation, following the reference workflow. For the main model, all minority categories were oversampled to match the largest group (84,362 samples per class, totalling 337,448 training records).

The predictive system integrates five separate sub-classifiers. We generated a stratified five-fold split of the training array, fitting each model on four folds while reserving the fifth. Predicted group probabilities for unseen entries are calculated by averaging outputs across all five sub-models.

##### Data Preprocessing

Products were retained only if they carried a valid NOVA label and non-missing values for all ten panel nutrients. Duplicate product codes were removed, keeping the first occurrence, and nutrient columns were coerced to numeric with non-numeric entries set to missing. Zero values were replaced with a small positive constant ($1\times10^{-20}$) and all nutrients were then natural-log transformed, following the original study's use of log-transformed nutrient features.

Before any class balancing or model fitting, the labelled Open Food Facts products with complete nutrient panels were split 80/20 into training and test partitions, stratified on NOVA class with a fixed random seed. Keeping a test set aside that the model never sees during training gives an honest estimate of how it performs on new foods. The 80/20 ratio is a common compromise between keeping enough data to train on and enough to test reliably; stratifying keeps the four NOVA groups in the same proportions in both parts, so that even the small NOVA 2 group (3.8%, 1,743 test products) is well represented; and the fixed seed means the same split can be reproduced exactly. This yielded 182,344 training and 45,586 test products. Splitting before balancing has two benefits: the test set keeps the database's natural class mix (21.90% NOVA 1, 3.82% NOVA 2, 27.90% NOVA 3, 46.37% NOVA 4), and it contains only real products, none of the artificial ones created during balancing (S3). Negative-value removal was then applied to each partition (208 training and 60 test products), leaving 182,136 training products (39,929 / 6,972 / 50,873 / 84,362 for NOVA 1–4) and 45,526 test products. The test partition was not examined until the final evaluation. All other analyses used Python 3.14 with scikit-learn 1.9.0, imbalanced-learn 0.14.2, SHAP 0.52.0, NumPy 2.4.6, and pandas 3.0.3.

#### S3. Training-data balancing and sensitivity model

Given that the primary authors utilized class-balanced training on Open Food Facts—a strategy frequently adopted in subsequent work—we fitted a secondary classifier where training instances were randomly down sampled to at most 7,000 items per category (27,972 products), and after removal of products with negative nutrient values 27,953 remained (7,000 / 6,972 / 6,999 / 6,982 for NOVA 1–4) before SMOTE was applied. To ensure direct comparability, performance for both predictive systems was assessed on the same held-out test split.

#### S4. Evaluation metrics and threshold-free UPF discrimination

The trained ensemble was evaluated, without any retraining or recalibration, on two databases with known NOVA labels: the Open Food Facts test partition held out before model fitting, and FNDDS 2009–10. Fineli carries no labels and is not part of the evaluation. Two complementary tasks were assessed: reconstruction of the full four-class NOVA taxonomy, and the binary task of identifying ultra-processed foods (NOVA 4) against all less-processed foods (NOVA 1–3).

Quantities derived from the predictions

For a given class, each food is one of four outcomes: a true positive (TP, correctly assigned to the class), a false positive (FP, wrongly assigned to the class), a false negative (FN, a true member of the class assigned elsewhere), or a true negative (TN, correctly kept out of the class):

- Precision = TP / (TP + FP) — of the foods the model places in the class, the proportion that truly belongs to it. Precision falls when the model over-assigns the class.
- Recall (sensitivity) = TP / (TP + FN) — of the foods that truly belong to the class, the proportion the model recovers. Recall falls when the model misses true members.
- F1 score = 2 · (precision · recall) / (precision + recall) — the harmonic mean of the two, which is high only when precision and recall are both high.
- Specificity = TN / (TN + FP) — the proportion of foods outside the class that the model correctly keeps out.
- Accuracy = the proportion of all foods assigned to their true class (for the four-class task, the trace of the confusion matrix divided by its total).

##### Four-Class Evaluation

For the four-class task we report overall accuracy, the full 4 × 4 confusion matrix, and precision, recall and F1 for each NOVA class computed one-versus-rest (that class as the positive label, the other three as the negative label).

##### Binary evaluation of ultra-processed foods

Because the question of practical interest is usually whether a food is ultra-processed, we additionally evaluated the binary task of separating NOVA 4 from NOVA 1–3. For this task, the model's score for a food is its predicted probability of NOVA 4, averaged over the five ensemble members. Turning that score into a yes/no call requires a decision threshold, and every threshold trades precision against recall: a high threshold flags only the foods the model is most sure about (high precision, low recall), a low threshold flags almost everything (high recall, low precision). Rather than committing to one threshold, we summarised performance across the whole range.

**Precision–recall curve and AUC-PR.** Sweeping the threshold from 1 to 0 traces a curve of precision (y-axis) against recall (x-axis). The area under this curve (AUC-PR; equivalently average precision, and the quantity of Menichetti et al. call AUP) condenses it into a single number between 0 and 1, with 1 being perfect and higher being better. AUC-PR does not have a fixed chance level: a non-informative classifier that assigns scores at random achieves an AUC-PR equal to the prevalence of the positive class — the fraction of foods that are ultra-processed. We therefore report AUC-PR alongside that prevalence baseline, so that the gain over chance is explicit; the same AUC-PR represents a larger gain in a database where ultra-processed foods are rarer.

**ROC curve and AUC-ROC.** Sweeping the same threshold and plotting recall against the false-positive rate (1 − specificity) gives the receiver operating characteristic curve. Its area (AUC-ROC) equals the probability that a randomly chosen ultra-processed food is given a higher NOVA-4 score than a randomly chosen food that is not ultra-processed; 0.5 is chance and 1 is perfect, regardless of prevalence. Because AUC-ROC is unaffected by class balance it is directly comparable between the Open Food Facts and FNDDS evaluations, which differ in the prevalence of ultra-processed foods; we report it for that comparison and AUC-PR as the more informative measure when the positive class is the minority.

#### S5. FNDDS zero-sentinel sensitivity analysis

Nutrient values in the FNDDS distribution are already log-transformed, and the original encoding represents log (0) as the sentinel value −20; that is, −20 denotes a nutrient that is absent rather than one that was not measured. Of the 2,971 FNDDS foods carrying a NOVA label in 1–4, 1,011 contained at least one such sentinel (2,026 values in total). In the primary analysis, these were interpreted as true zeros and re-expressed in the zero-encoding used during training, retaining all 2,971 foods. As a sensitivity analysis, we instead treated them as missing and excluded the affected foods, leaving 1,960. The former is preferred: excluding them removes a third of the labelled foods, biases the evaluation set toward nutrient-complete products, and reduces NOVA 2 support from 54 foods to 3.

#### S6. Original FoodProX comparison

Two comparisons were made against the classifier released by Menichetti et al. 2023 On FNDDS, where their model's predicted probabilities are distributed with the database, we computed the agreement between their predictions and ours on identical foods; their accuracy on these foods is an in-sample figure, as FNDDS 2009–10 is the database on which their model was trained, and is reported as such rather than as a comparable out-of-sample benchmark.

On Fineli, where no labels exist, we applied their released twelve-nutrient classifier directly and compared it with ours on the same foods, reporting class agreement and the rank correlation of the two FPro scores. Their panel differs from ours: it comprises protein, total fat, carbohydrate, total sugars, dietary fibre, calcium, iron, sodium, vitamin C, cholesterol, saturated fat and total vitamin A, contains no energy term, and expresses all nutrients in grams per 100 g. Fineli values were rescaled accordingly (mg to g for minerals, vitamin C and cholesterol; µg to g for vitamin A). Their released models cannot be loaded by scikit-learn 1.3 or later, so we ran them in a separate environment (Python 3.8, scikit-learn 0.23.2) and passed their predictions to the main analysis as a plain CSV file.

#### S7. FPro calculation and uncertainty

The food processing score (FPro) is a continuous variable that captures the discrete classes of NOVA on a continuum. For each food sample $k$, FPro is defined following Menichetti et al. (2023) as:

$$\mathrm{FPro}_{k,f} = \frac{1 - p_1^{k,f} + p_4^{k,f}}{2}, \qquad f = 1, \dots, 5 \qquad (7.1).$$

where $p_1^{k,f}$ and $p_4^{k,f}$ are the predicted probabilities of NOVA 1 and NOVA 4 per classifier $f$. Because each food is scored by all five ensemble classifiers, $FPro_k$ is computed once per classifier $f$ using Eq. S7.1 (giving $FPro_{k,f}, f = 1, \dots, 5$) before aggregating. The reported score is the mean across the five classifiers, and its uncertainty is the sample standard deviation across them:

$$\overline{\mathrm{FPro}}_k = \frac{1}{5}\sum_{f=1}^{5} \mathrm{FPro}_{k,f}\ , \qquad \sigma_{\mathrm{FPro},k} = \sqrt{\frac{1}{5-1}\sum_{f=1}^{5}\left(\mathrm{FPro}_{k,f} - \overline{\mathrm{FPro}}_k\right)^2} \qquad (7.2).$$

FPro ranges from 0 (unprocessed) to 1 (ultra-processed). We also report $min_f(\mathrm{FPro}_{k,f})$ as a conservative bound, following the original implementation. Computing the score per classifier before aggregating, rather than propagating the variances of $p_1$ and $p_4$ under an assumption of independence, is necessary because the two probabilities are negatively correlated across ensemble members; the independence approximation understates SD(FPro).

##### S8. Permutation importance and SHAP analysis

To identify which nutrients, drive the classification we computed permutation importance on the Open Food Facts test partition, measuring the loss in accuracy when each nutrient is randomly shuffled, on 20,000 held-out test-set products with 5 repeats per nutrient, for both model configurations. We additionally computed SHAP values (TreeExplainer) summarised as mean absolute SHAP per nutrient per NOVA class, on 2,000 test-set foods for the sensitivity (class-balanced) configuration, across all five ensemble members; this configuration was used because the primary forest is roughly twelve times larger, and permutation importance ranks the same two nutrients first in both configurations.

### Supplementary Results

##### SR1. Cross-validation and held-out performance

###### Classifier Performance in the training database

Five-fold cross-validation on the Open Food Facts training partition yielded high discrimination for all four NOVA classes (AUC 0.996, 0.999, 0.972 and 0.976 for NOVA 1-4; overall accuracy 0.911 ± 0.001; Table S1). These cross-validated values are optimistic. To balance the classes, SMOTE creates artificial examples of the rarer NOVA groups by blending pairs of similar real foods. As in the original implementation, this was done before the training data were divided into five parts for cross-validation, so 46.0% of the rows (155,312 of 337,448) were artificial, and many were blended from foods that ended up in the part used for testing. The model was therefore partly tested on foods resembling ones it had already seen, which inflates the estimate. Accuracy on the untouched held-out test set (0.855) is the realistic figure; part of the difference also reflects that set's natural class mix (see Discussion). In plain terms, part of each test fifth was not truly unseen, so the estimate is flattering.

**Table S1**

| | AUC1 | AUC2 | AUC3 | AUC4 | CV acc. | CV acc. SD |
|---|---|---|---|---|---|---|
| Full SMOTE (Primary) | 0.9959 | 0.9993 | 0.9717 | 0.9758 | 0.9113 | 0.0012 |
| Balanced (Sensitivity) | 0.9866 | 0.9941 | 0.9303 | 0.9422 | 0.8426 | 0.0041 |

**Table S1.** Five-fold stratified cross-validation of the two model configurations. Per-class ROC-AUC (one-vs-rest, NOVA 1–4) averaged across folds, and cross-validated accuracy with its standard deviation across folds. Primary: full training set with SMOTE; sensitivity: classes undersampled to 7,000 products each. SMOTE was applied before the folds were drawn, as in the original FoodProX code, so these estimates are optimistic; held-out test performance (accuracy 0.855) is reported in Table S2.

#### SR2. External validation in FNDDS

##### Cross-national transfer to the United States food supply

Applied without retraining or recalibration to FNDDS 2009–10, the classifier achieved an overall accuracy of 0.763 on 2,971 labelled foods, against 0.855 on the held-out European data, a loss of about nine percentage points. Threshold-free discrimination of ultra-processed foods degraded more modestly (AUC-ROC 0.951 → 0.919).

The degradation was concentrated in the intermediate classes rather than distributed evenly (Figure S4a). Recall for NOVA 4 fell only from 0.86 to 0.82 and for NOVA 1 from 0.95 to 0.88, whereas recall for NOVA 3 fell from 0.76 to 0.44 (NOVA 2: 0.95 to 0.69, on 54 foods). F1 for NOVA 1 nevertheless fell from 0.92 to 0.65, because precision dropped from 0.90 to 0.52 as US NOVA 3 and NOVA 4 foods were assigned to NOVA 1 (Table S4; Figure S4). The model's ability to recover NOVA 1 and NOVA 4 foods therefore appears largely conserved between the European and United States food supplies, while the intermediate category transfers poorly and NOVA 1 predictions become less reliable.

Transfer also introduced a second, distinct error mode. Within Open Food Facts most errors (74%) were between adjacent classes; in FNDDS, 20.6% of errors were true NOVA 3 foods called NOVA 1 and a further 17.9% were true NOVA 4 foods called NOVA 1, so that the NOVA 3 ↔ 4 confusion fell to 48.4% of errors. The model does not merely lose resolution under transfer: it systematically under-calls the degree of processing of United States foods, consistent with the lower mean FPro it assigns relative to the original classifier on the same foods (0.678 versus 0.774; Table S5).

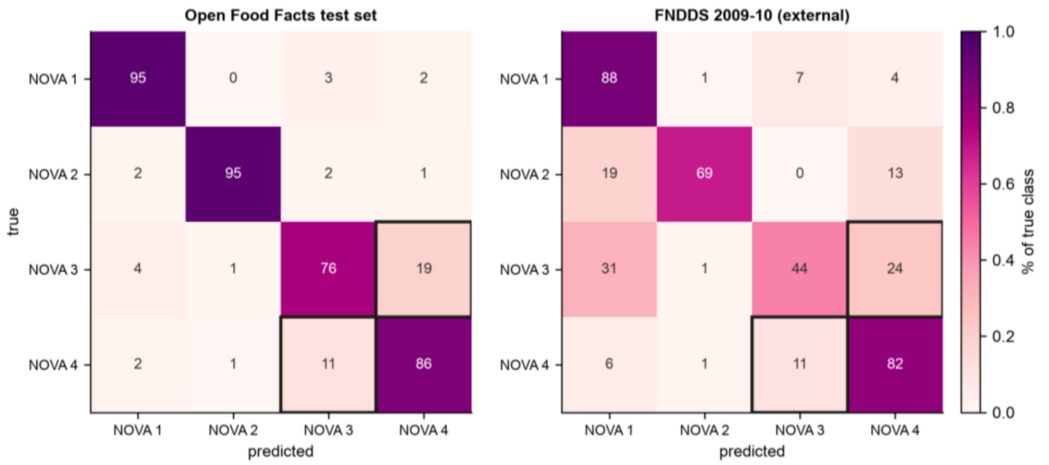


**Figure S4. Confusion matrices.** Row-normalised four-class confusion matrices (percentages of each true class) for the primary configuration on the Open Food Facts test set and on FNDDS; the NOVA 3 ↔ 4 cells are outlined. Under transfer, true NOVA 3 and NOVA 4 foods leak into the NOVA 1 column —an error mode about a quarter as common within Europe (2.3% vs 9.1% of foods) — indicating systematic under-calling of processing in the United States food supply.

#### Table S2

| **configuration** | **evaluation** | **accuracy** | **$F1_1$** | **$F1_2$** | **$F1_3$** | **$F1_4$** |
|---|---|---|---|---|---|---|
| Primary (full data + SMOTE) | OFF test set | 0.855 | 0.92 | 0.91 | 0.77 | 0.87 |
| Primary | FNDDS (zero, primary) | 0.763 | 0.65 | 0.58 | 0.44 | 0.87 |
| Primary | FNDDS (missing, sensitivity) | 0.788 | 0.69 | 0.33 | 0.39 | 0.89 |
| Sensitivity (class-balanced) | OFF test set | 0.799 | 0.89 | 0.87 | 0.70 | 0.81 |
| Sensitivity | FNDDS (zero) | 0.742 | 0.66 | 0.51 | 0.48 | 0.85 |
| Sensitivity | FNDDS (missing) | 0.768 | 0.71 | 0.22 | 0.42 | 0.87 |

**Table S2. Classification accuracy and per-class F1.** Overall accuracy and per-class F1 score for both model configurations on the Open Food Facts test set and on FNDDS under both sentinel policies.

##### Table S3

| configuration | evaluation | n | UPF_prevalence | AUC_PR | AUC_PR_baseline | AUC_ROC |
|---|---|---|---|---|---|---|
| Primary (full training data + SMOTE) | OFF test set | 45526 | 0.463 | 0.9399 | 0.463 | 0.9513 |
| Primary (full training data + SMOTE) | FNDDS (zero) | 2971 | 0.7109 | 0.9635 | 0.7109 | 0.9191 |
| Primary (full training data + SMOTE) | FNDDS (missing) | 1960 | 0.7454 | 0.9783 | 0.7454 | 0.9448 |
| Sensitivity (class-balanced undersampling) | OFF test set | 45526 | 0.463 | 0.8988 | 0.463 | 0.9182 |
| Sensitivity (class-balanced undersampling) | FNDDS (zero) | 2971 | 0.7109 | 0.9606 | 0.7109 | 0.9132 |
| Sensitivity (class-balanced undersampling) | FNDDS (missing) | 1960 | 0.7454 | 0.9768 | 0.7454 | 0.9424 |

**Table S3. Threshold-free discrimination of ultra-processed foods**. For each model configuration and evaluation set: the number of foods, the prevalence of ultra-processed foods, the AUC-PR, its prevalence baseline (the AUC-PR of a non-informative classifier), and the AUC-ROC. AUC-PR must be read against its baseline, which differs between databases. OFF (Open Food facts)

##### Table S4

| configuration | evaluation | prec_1 | rec_1 | prec_2 | rec_2 | prec_3 | rec_3 | prec_4 | rec_4 |
|---|---|---|---|---|---|---|---|---|---|
| Primary (full training data + SMOTE) | OFF test set | 0.898 | 0.948 | 0.873 | 0.953 | 0.784 | 0.758 | 0.873 | 0.861 |
| Primary (full training data + SMOTE) | FNDDS (zero, primary) | 0.516 | 0.882 | 0.5 | 0.685 | 0.447 | 0.442 | 0.93 | 0.817 |
| Primary (full training data + SMOTE) | FNDDS (missing, sensitivity) | 0.556 | 0.919 | 0.333 | 0.333 | 0.376 | 0.41 | 0.951 | 0.84 |
| Sensitivity (class-balanced undersampling) | OFF test set | 0.847 | 0.942 | 0.788 | 0.967 | 0.684 | 0.72 | 0.854 | 0.766 |
| Sensitivity (class-balanced undersampling) | FNDDS (zero) | 0.517 | 0.906 | 0.404 | 0.704 | 0.426 | 0.541 | 0.95 | 0.761 |
| Sensitivity (class-balanced undersampling) | FNDDS (missing) | 0.568 | 0.942 | 0.167 | 0.333 | 0.355 | 0.502 | 0.967 | 0.793 |

**Table S4. Per-class precision and recall.** Precision and recall for each NOVA class, for both model configurations, on the Open Food Facts test set and on FNDDS under both sentinel policies.

#### SR3. Agreement with the original released classifier

**Table S5**— Our model vs. the original Menichetti et al. released classifier, both scored on the same FNDDS foods (the US database), under the two sentinel policies (−20 read as zero vs. as missing).

| Evaluation | n compared | 4-class agreement | UPF-vs-rest agreement | FPro Spearman | FPro Pearson | Our accuracy (out-of-sample) | Their accuracy (in-sample) | Mean FPro (ours) | Mean FPro (theirs) |
|---|---|---|---|---|---|---|---|---|---|
| FNDDS (zero) | 2,971 | 0.765 | 0.829 | 0.813 | 0.809 | 0.763 | 0.996 | 0.678 | 0.774 |
| FNDDS (missing) | 1,960 | 0.790 | 0.851 | 0.811 | 0.860 | 0.788 | 0.998 | 0.699 | 0.795 |

**Table S5. FNDDS: This study versus the original released classifier.** For each sentinel policy: the number of foods compared, four-class and ultra-processed / not agreement between the two models, Spearman and Pearson correlation of the FPro scores, our out-of-sample accuracy, and the original model's accuracy on the same foods. The latter is an in-sample figure — FNDDS 2009–10 is the database on which that model was trained — and is not a comparable benchmark.

**Table S6** — Our model vs. the original Menichetti et al. released classifier, applied side by side to Fineli (the Finnish database, which has no NOVA labels, so this is agreement, not accuracy).

| n foods compared | 4-class agreement | UPF-vs-rest agreement | FPro Spearman | FPro Pearson | Mean FPro (ours) | Mean FPro (original) | FPro bias (ours − original) |
|---|---|---|---|---|---|---|---|
| 3,892 | 0.793 | 0.828 | 0.789 | 0.878 | 0.642 | 0.704 | −0.062 |

**Table S6. Fineli: This study versus the original released classifier.** On the 3,892 Fineli foods scorable by both nutrient panels: four-class agreement, ultra-processed / not agreement, Spearman and Pearson correlation of the two FPro scores, the mean FPro of each model, and the mean difference. Neither model has ground truth on Fineli; agreement is convergent validity, not accuracy.

#### SR4. Supplementary evidence on nutrients effect on classification

SHAP values (TreeExplainer; 2,000 held-out foods; all five ensemble members; Figure S5) agree that sodium matters most: it carries the largest mean absolute attribution (0.095), with protein (0.051) and sugars (0.050) close together behind it. They also resolve which nutrient carries which class (Figure S5a). Attribution is strongly class-specific: sodium dominates the NOVA 1 (0.162) and NOVA 3 (0.112) probabilities; protein dominates NOVA 2 (0.084), and total sugars dominate NOVA 4 (0.082), the class of practical interest. The model thus recognises unprocessed foods by their low sodium, processed foods by their high sodium, culinary ingredients by their near absence of protein, and ultra-processed foods primarily by their sugar. The attributions are also monotone in the nutrient itself (Figure S5b): across the 2,000 explained foods, the rank correlation between a nutrient's level and its SHAP value for the NOVA 4 probability was +0.87 for sugars and +0.84 for sodium, and −0.59 for iron and −0.57 for dietary fibre: sugar and salt push a food towards the ultra-processed pole while fibre and iron, markers of whole-food matrices, pull it away. Saturated fat contributed ≤ 0.030 in every class.

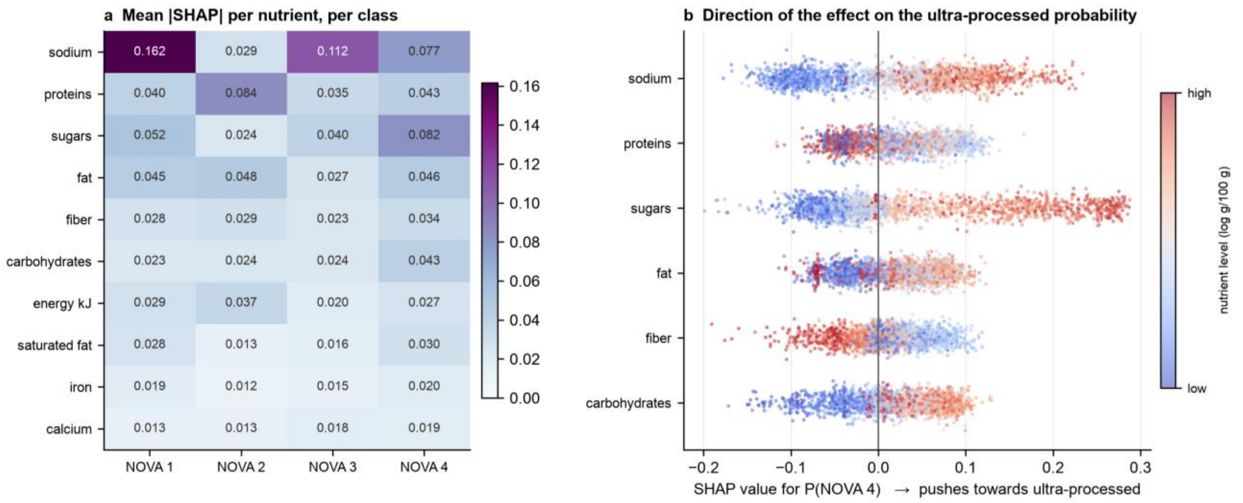


**Figure S5. Nutrient attribution by SHAP.** Computed with TreeExplainer on 2,000 test-set foods for the sensitivity configuration (all five ensemble members). (a) Mean absolute SHAP value per nutrient per NOVA class. Attribution is class-specific: sodium dominates the NOVA 1 and NOVA 3 probabilities, protein dominates NOVA 2, and total sugars dominate NOVA 4. (b) SHAP values for the probability of NOVA 4 across the 2,000 foods, for the six highest-attribution nutrients, with points colored by the food's rank on that nutrient (blue low, red high). High sodium and high sugar push food towards the ultra-processed pole; high fibre pulls it away.

The SHAP analysis was run on the class-balanced (sensitivity) model because the primary forest is roughly twelve times larger; permutation importance ranks the same two nutrients first in that model as in the primary one (sodium 21.8%, sugars 12.3% of baseline accuracy).

#### Table S7

| nutrient | Accuracy drop (primary) | % of baseline (primary) | Accuracy drop (sensitivity) | % of baseline (sensitivity) |
|---|---|---|---|---|
| sodium_100g | 0.1549 | 18.1177 | 0.1725 | 21.7941 |
| sugars_100g | 0.0864 | 10.1053 | 0.0975 | 12.3184 |
| proteins_100g | 0.0614 | 7.1888 | 0.0395 | 4.9905 |
| carbohydrates_100g | 0.0446 | 5.2129 | 0.0332 | 4.2009 |
| fiber_100g | 0.0403 | 4.7181 | 0.0348 | 4.3904 |
| calcium_100g | 0.036 | 4.215 | 0.0252 | 3.1901 |
| iron_100g | 0.0345 | 4.0314 | 0.0357 | 4.5167 |
| fat_100g | 0.0343 | 4.0173 | 0.0302 | 3.8219 |
| energy-kj_100g | 0.0331 | 3.8711 | 0.0308 | 3.885 |
| saturated fat_100g | 0.0204 | 2.3854 | 0.0162 | 2.0531 |

**Table S7. Permutation importance of each nutrient**: the fall in four-class accuracy on the held-out test set when that nutrient's values are randomly shuffled, as an absolute drop and as a percentage of unshuffled accuracy. Primary configuration: 20,000 products, 5 shuffles, baseline accuracy 0.855. Sensitivity configuration: 2,000 products, 2 shuffles, baseline 0.792 (0.799 on the full test set, Table S2). Rows are ordered by importance in the primary configuration.

**Table S8**

| Nutrient | Spearman ρ (NOVA 4) | p-value | n |
|---|---|---|---|
| **Total sugars** | 0.869 | <1e-100 | 2,000 |
| **Sodium** | 0.837 | <1e-100 | 2,000 |
| **Carbohydrate** | 0.696 | <1e-100 | 2,000 |
| **Iron** | −0.591 | <1e-100 | 2,000 |
| **Saturated fat** | 0.583 | <1e-100 | 2,000 |
| **Dietary fibre** | −0.574 | <1e-100 | 2,000 |
| **Total fat** | 0.356 | 5.62e-61 | 2,000 |
| **Calcium** | −0.273 | 2.17e-35 | 2,000 |
| **Protein** | −0.116 | 2.09e-07 | 2,000 |
| **Energy (kJ)** | −0.111 | 6.18e-07 | 2,000 |

**Table S8. Rank correlation between nutrient level and SHAP attribution for the ultra-processed (NOVA 4) probability.** Companion to Figure S5b: the beeswarm shows this relationship visually but carries no printed value, so this table gives the number behind it. For each nutrient, Spearman's ρ between its raw level and its SHAP value for the NOVA 4 probability, computed on the same 2,000 test-set foods and the same sensitivity-configuration SHAP values as Figure S5 (TreeExplainer, all five ensemble members). Positive ρ means higher levels of that nutrient push a food predicted probability towards ultra-processed; negative ρ means higher levels pull it away. Columns: **nutrient**; **spearman_rho_NOVA4**; **p_value**;**n** — food entering the correlation (2,000 throughout). Rows are ordered by |ρ|. Total sugars (ρ = 0.869) and sodium (ρ = 0.837) show the strongest positive association; iron (ρ = −0.591) and dietary fibre (ρ = −0.574) the strongest negative one — consistent with the sodium–sugar axis identified in Table S7 and the class-specific attribution in Figure S5a.

#### SR5. Probability Space and uncertainty of model output on FINELI

**Probability geometry.** Projected onto the first two principal components of the four predicted probabilities (Figure S6), Finnish foods occupy the corners of the probability simplex corresponding to confident NOVA 1, 3 and 4 assignments, with a populated interior in which no class dominates. This is the same geomyetry as the original study reported for FNDDS and is the structural reason a continuous score is needed: for interior foods the assignment of a single NOVA class is close to arbitrary, whereas FPro orders them smoothly.

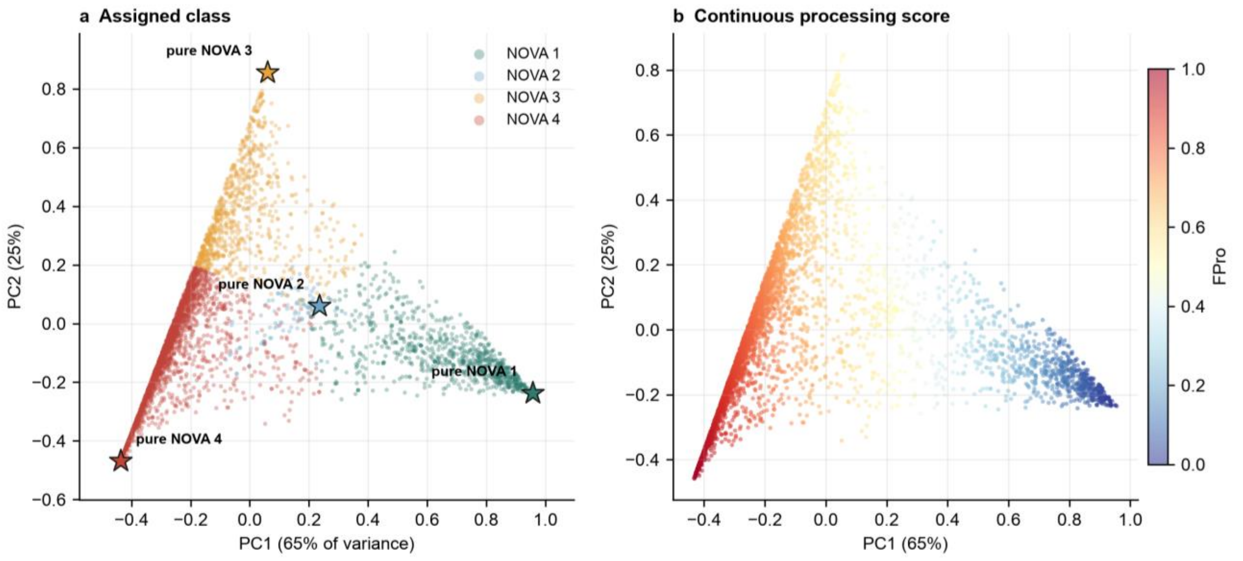


**Figure S6. Fineli in the classifier's probability space.** Principal-component projection of the four predicted class probabilities for the 4,092 scored Fineli items. (a) Coloured by assigned NOVA class; stars mark the projected pure-class vertices. Foods with a dominant class sit near the vertices; the populated interior contains foods for which a single NOVA label is close to arbitrary. (b) The same points coloured by FPro, which orders the interior smoothly. This reproduces the decision-space geometry the original study reported for FNDDS.

**Uncertainty.** Ensemble uncertainty was low for most items: the mean standard deviation of FPro across the five classifiers was 0.015, and 80.4% of items had σ_FPro < 0.02 (Figure S7). Uncertainty was lowest at the two poles and highest for foods scoring between roughly 0.2 and 0.5, again locating the model's indecision in the middle of the processing range (NOVA 2-3). σ_FPro is released per food so downstream users can threshold on it.

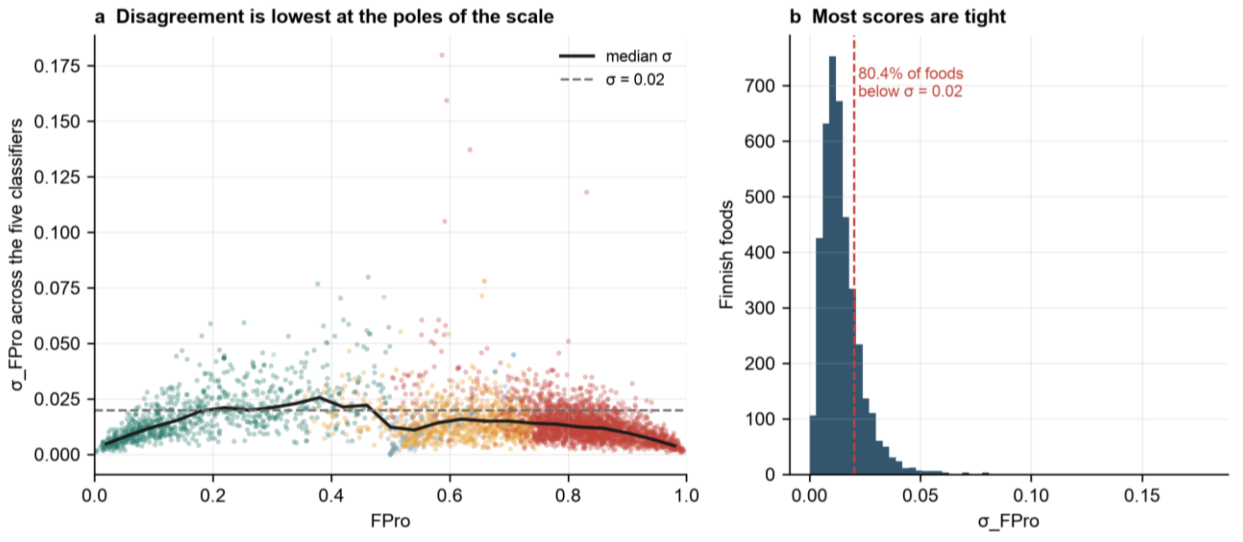


**Figure S7. Per-food confidence in the Fineli annotation.** (a) σ_FPro against FPro for the 4,092 scored items, with a running median and the σ = 0.02 reference line; ensemble disagreement is lowest at the two poles and peaks for foods scoring roughly 0.2–0.5. (b) Distribution of σ_FPro: 80.4% of items fall below 0.02.

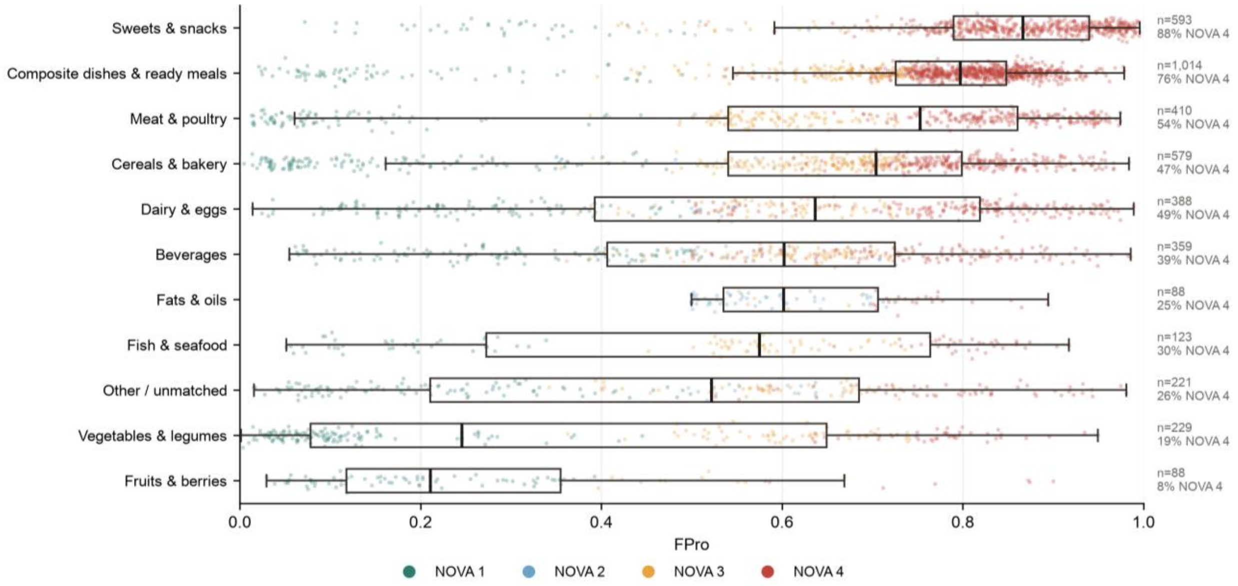


**Figure S8. Fineli FPro by food group.** Box plots with jittered points of FPro for eleven name-derived food groups, ordered by median, with per-group n and the percentage assigned NOVA 4. The ordering is the expected one (fruits and vegetables lowest, sweets and composite dishes highest); the spread within each group is wide, which is the point — FPro separates foods that both NOVA and food-group taxonomies collapse. Groups are descriptive, derived from item names, and are not an official Fineli classification.

#### SR6. Sensitivity analyses

Class balance. Randomly undersampling the training partition to 7,000 products per class before SMOTE, a common default in this literature, reduced held-out accuracy from 0.855 to 0.799, FNDDS accuracy from 0.763 to 0.742, and binary AUC-PR from 0.940 to 0.899 (Table S2, Table S3). Discarding roughly 85% of the training data to balance classes was costly, so the full-data model is reported as primary.

FNDDS missing-value sentinel. Interpreting the −20 sentinel as "not measured" and excluding the affected foods raised apparent accuracy from 0.763 to 0.788 but did so by discarding a third of the labelled foods and reducing NOVA 2 support from 54 to 3, making per-class estimates for that group uninterpretable. The higher figure is an artefact of a more nutrient-complete, and therefore easier, evaluation set.

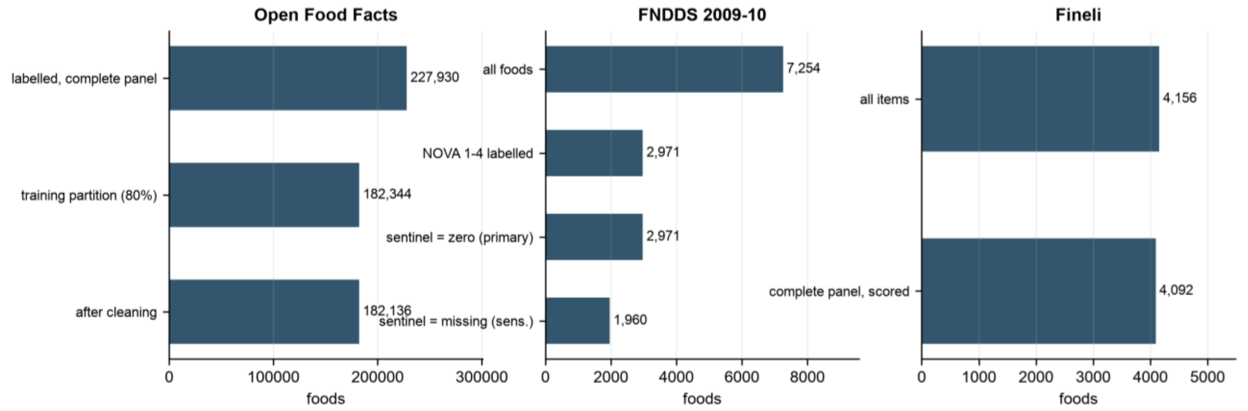


**Figure S9. Foods are retained at each stage.** Counts remaining after each cleaning and filtering step, for Open Food Facts, FNDDS and Fineli. For FNDDS the choice between reading the −20 sentinel as a true zero (retained) or as missing (dropped) is the difference between 2,971 and 1,960 labelled foods, and between 54 and 3 NOVA 2 foods.
